# Competing Risks in Patients with Primary Prevention Implantable Cardioverters Defibrillators: Global Electrical Heterogeneity and Clinical Outcomes (GEHCO) Study

**DOI:** 10.1101/2020.12.16.20248369

**Authors:** Jonathan W. Waks, Kazi T. Haq, Christine Tompkins, Albert J. Rogers, Ashkan Ehdaie, Aron Bender, Jessica Minnier, Khidir Dalouk, Stacey Howell, Achille Peiris, Merritt Raitt, Sanjiv M. Narayan, Sumeet S. Chugh, Larisa G. Tereshchenko

## Abstract

**Background:** Global electrical heterogeneity (GEH) is associated with sudden cardiac death in the general population.

**Objective:** To investigate whether GEH is associated with sustained ventricular tachycardia (VT)/ventricular fibrillation (VF) leading to appropriate implantable cardioverter-defibrillator (ICD) therapies in systolic heart failure (HF) patients with primary prevention ICDs.

**Methods:** We conducted a multicenter retrospective cohort study. GEH was measured by spatial ventricular gradient (SVG) direction and magnitude, QRS-T angle, and sum absolute QRST integral (SAIQRST) on pre-implant 12-lead ECGs. Survival analysis using cause-specific hazard functions compared the strength of associations with two competing outcomes: sustained VT/VF leading to appropriate ICD therapies and all-cause death without appropriate ICD therapies.

**Results:** We analyzed data from 2,668 patients (age 63±12y; 23% female; 78% white; 43% nonischemic cardiomyopathy (NICM); left ventricular ejection fraction 28±11% from 6 academic medical centers). After adjustment for demographic, clinical, device, and traditional ECG characteristics, SVG elevation (Hazard Ratio (HR) per 1 standard deviation (SD) 1.14 (95% CI 1.04-1.25); *P*=0.004), SVG azimuth (HR 1.12(1.01-1.24); *P*=0.039); SVG magnitude (HR per 1 SD 0.75 (0.66-0.85); *P*<0.0001), and QRS-T angle (HR per 1 SD 1.21 (95% CI 1.08-1.36); *P*=0.001) were associated with appropriate ICD therapies. The SVG azimuth was also associated with a lower competing risk of death (*P*_*difference*_=0.007): HR 0.91(0.82-1.02); *P*=0.095. SAIQRST had different associations in ischemic [HR 1.29(1.04-1.60)] and NICM [HR 0.78(0.62-0.96); *P*_*interaction*_=0.022].

**Conclusion:** In patients with primary prevention ICDs, GEH is independently associated with appropriate ICD therapies. The SVG vector points in distinctly different directions in patients with two competing outcomes.

**Clinical Trial Registration:** URL:www.clinicaltrials.gov Unique identifier: NCT03210883.

## Introduction

Patients with cardiomyopathy (CM) and reduced left ventricular ejection fraction (LVEF) are at elevated risk of sudden cardiac death (SCD), and in this population, current guidelines recommend consideration of implantable cardioverter-defibrillators (ICDs) for primary prevention of SCD^1^. However, current guidelines for implantation of primary prevention ICDs rely primarily on LVEF and the New York Heart Association (NYHA) functional class, which are overall poor predictors of ICD benefit, especially in patients with nonischemic CM (NICM).^2^ As ICD implantation is associated with significant costs for the medical system and risk to individual patients, improved methods of identifying patients most likely to benefit from primary prevention ICDs are needed.

Global electrical heterogeneity (GEH)^3, 4^ is a comprehensive characterization of the spatial ventricular gradient (SVG) vector^5^ by its 5 features: SVG vector direction (azimuth and elevation), magnitude, its scalar value, and spatial QRS-T angle.^3^ It integrates static and dynamic alternations in ventricular repolarization and conduction, which have been linked to ventricular arrhythmias in studies from the bench to the bedside.^6^ Abnormal GEH is associated with SCD in the general population^3, 7^ where it is explicitly associated with SCD over non-arrhythmic modes of death^3^, and an increased risk of drug-induced torsades-de-pointes.^8, 9^ The utility of assessing GEH for SCD risk stratification in CM patients with reduced LVEF who meet primary prevention ICD indications, however, is unknown. As GEH might be a useful way of identifying patients who are most (or least) likely to benefit from primary prevention ICD implantation, we conducted a novel retrospective multicenter cohort study^10^ to determine the associations of the clinical, device, traditional ECG, and GEH metrics with sustained ventricular arrhythmias requiring appropriate ICD therapies and competing death without appropriate ICD therapies.

## Methods

The multicenter study has been approved by the Oregon Health & Science University (OHSU) Institutional Review Board (IRB). In addition, each participating center obtained local IRB approval prior to participating.

### Study design

The study design and protocol has been previously described.^10^ In brief, we conducted a retrospective, multicenter cohort study that included data from six academic medical centers in the United States: OHSU in Portland, OR, Veteran Administration Portland Healthcare System (Portland VA) in Portland, OR, Beth Israel Deaconess Medical Center (BIDMC) in Boston, MA, the University of Colorado (U Colorado) in Aurora, CO, Cedars-Sinai Medical Center (CSMC) in Los Angeles, CA, and Stanford University (Stanford U) in Palo Alto, CA. We included all patients above the age of 18 years with chronic systolic heart failure (HF) due to infarct-related CM and/or NICM, who underwent implantation of any ICD [including single chamber, dual-chamber, cardiac resynchronization therapy defibrillator (CRT-D), or subcutaneous] for primary prevention of SCD between the years of 1996 and 2019, and had an available digital 12-lead ECG recording. Patients with inherited channelopathies and cardiomyopathies, congenital heart disease, and those with ICD implanted for secondary prevention were excluded.

### ECG analysis: global electrical heterogeneity measurement

A digital 12-lead ECG recorded around the time of initial ICD implantation was collected (median 0 days (interquartile range 0-29 days). Raw, digital ECG signals were analyzed in the Tereshchenko laboratory at OHSU, as previously described.^3, 7, 11, 12^ At least two physician-investigators manually labeled each cardiac beat (SH, EB, LGT). The Kors transformation matrix was used to transform 12-lead ECGs into orthogonal XYZ vectorcardiograms.^13^ The time-coherent global median beat was constructed using the dominant type of beat present in the ECG, and the origin point of the vectorcardiogram was identified.^12^ We included three categories of median beats: normal (N) median beats included normal sinus rhythm, atrial pacing, junctional rhythm, and ectopic atrial rhythm; ventricular paced (VP) median beats included right ventricular or biventricular pacing; supraventricular (S) median beats included atrial fibrillation or atrial flutter with a consistent QRS morphology. QT interval was corrected using Bazett, Fridericia, and Hodges equations. The presence of premature ventricular complexes (PVCs) on 12-lead ECG was noted.

We constructed spatial peak and area QRS and T vectors as well as their vector sum (SVG) and measured their magnitude, direction (azimuth (orientation in the XZ plane) and elevation (orientation in the XY plane)), and spatial QRS-T angles (Figure 1).^7, 11, 12^ Scalar approximations of the SVG were measured via the sum absolute QRST integral (SAIQRST)^14-16^ and the QT integral on vector magnitude (VMQTi).^11^ Quality control of automated ECG analysis was performed to verify appropriate beat identification and fiducial point annotation (KTH). Open-source MATLAB (MathWorks, Natick, MA, USA) code for GEH measurement is provided at https://physionet.org/physiotools/geh & https://github.com/Tereshchenkolab/Origin.

**Figure 1.**
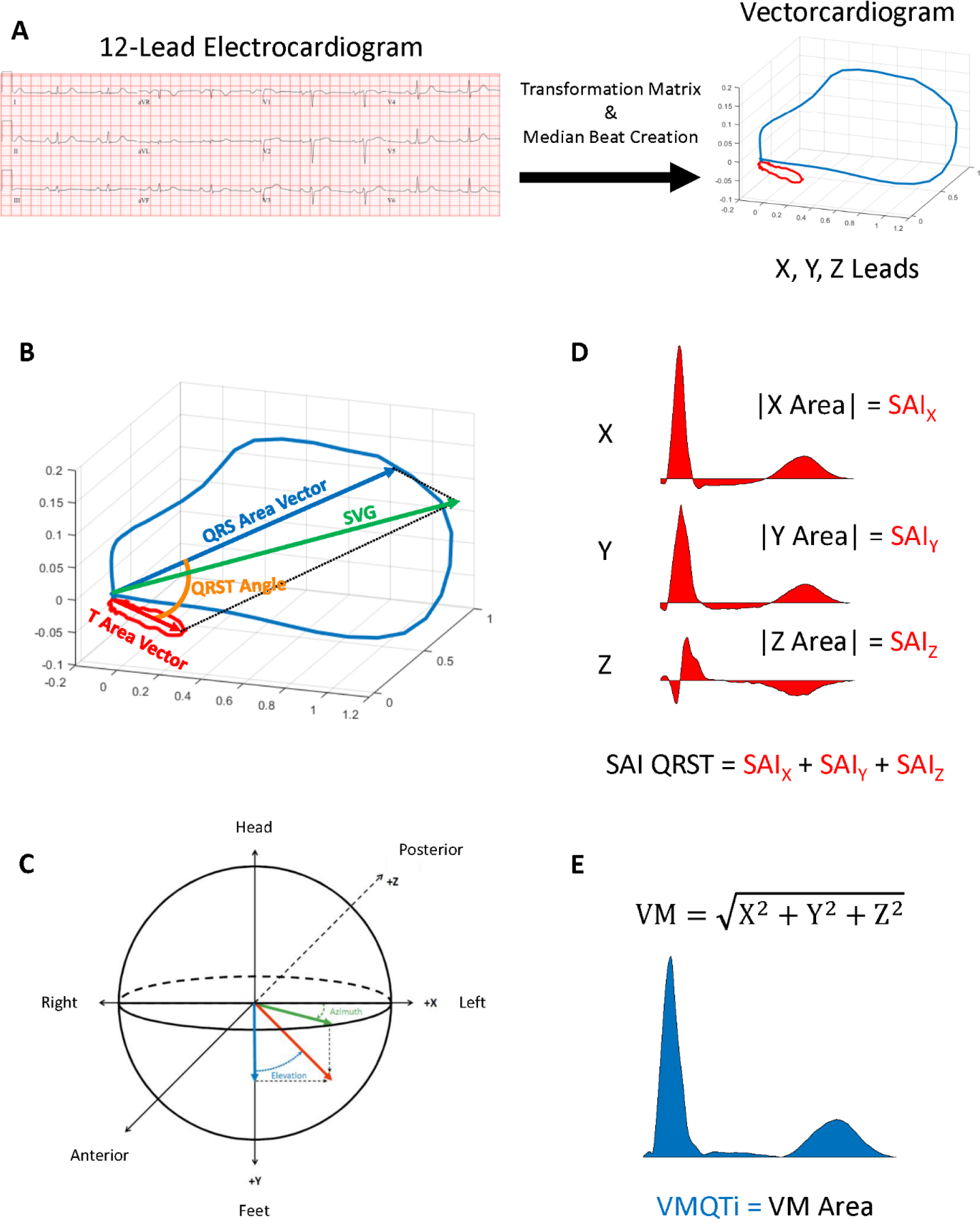
GEG measurements. **A**. Transformation of a 12-lead ECG to orthogonal XYZ ECG. **B**. The spatial ventricular gradient (SVG) is a vector formed from the sum of the area QRS and area T wave vectors. The QRST angle is the 3-dimensional angle between the area QRS and area T wave vectors. **C**. Orientation of angles in 3D space. Positive angles are posterior and negative angles are anterior. **D**. SAIQRST is the sum of the absolute areas of the X, Y, and Z leads. **E**. The vector magnitude lead (VM) is calculated as VM=√(X^2^+Y^2^+Z^2^). The VM QT integral (VMQTi) is the area under the QT ECG signal on VM lead.

### Primary Competing Outcomes

The primary outcome was defined as the first sustained ventricular tachyarrhythmia event (VT or VF) with appropriate ICD therapy (either anti-tachycardia pacing or shock). All-cause death without preceding appropriate ICD therapy served as the primary competing outcome.

### Clinical characteristics and ICD programming

Patients’ demographics, clinical history, and laboratory values at the pre-implant clinical evaluation were abstracted from the medical record. Estimated glomerular filtration rate (eGFR) was calculated using the Chronic Kidney Disease Epidemiology Collaboration equation.^17^ ICD detection parameters were obtained at the time of device implant or at the time of appropriate ICD therapies if they occurred during follow-up. To harmonize VT/VF detection duration programming across different device manufacturers, we grouped patients^18^ in (1) long detection duration (number of intervals to detect (NID) VT ≥ 30/40 or VT detection duration ≥ 10s or VF detection duration ≥ 3s) or (2) short detection duration (NID VT <30/40 or VT detection duration < 10s or VF detection duration < 3s) categories (Supplemental Table 1).

### Statistical analyses

Normally distributed continuous variables were reported as means and standard deviation (SD). Circular variables were reported as mean and 95% confidence interval (CI). An unadjusted comparison of circular variables (spatial QRS-T angle, SVG azimuth, and SVG elevation) was performed using the Wheeler-Watson-Mardia test. ANOVA (for normally distributed continuous variables) and Pearson’s χ^2^ test (for categorical variables) were used to compare ECG characteristics in patients with and without primary competing outcomes. Unadjusted analyses were performed using the dataset with missing values.

As we conducted a retrospective study, the data contained missing values for reasons unrelated to the study itself. Missingness data for individual covariates are reported in Supplemental Table 2. The most likely reason for missing values (incomplete data in the medical record) was unrelated to the unobserved data but likely was dependent upon the observed data (e.g., ICD therapy), which is characteristic for data missing at random. Therefore, we employed multiple imputations using chained equations (MICE)^19^, which does not require the multivariate normal assumption. Our study’s investigation of data missingness showed an arbitrary pattern. Observations with missing primary outcomes were excluded. We used predictive mean matching with 20 nearest neighbors for continuous variables, logistic regression for binary variables, ordinal logistic regression for ordinal variables, and multinomial logistic regression for nominal variables with 66 imputations, exceeding the minimum required by the von Hippel method.^20^ Convergence was confirmed. Relative variance increase, fraction missing information, and relative efficiency were assessed for each multivariable model and confirmed acceptable.

After examining a normal quantile plot, we included QRS-T and SVG elevation angles in all adjusted statistical analyses without transformation because their distributions were nearly normal. The SVG azimuth angle is ranging from -180° to +180°. As recommended,^21-23^ we transformed SVG azimuth by doubling its value and then adding 360°.

As the risk of sustained VT/VF leading to appropriate ICD therapy competes with the risk of death without appropriate ICD therapy, we employed cause-specific hazards functions, estimated using Cox proportional hazards models. For a model with sustained VT/VF and appropriate ICD therapy outcome, all-cause death event was censored at the date of death if there was no documented sustained ventricular tachyarrhythmia at any time since implant until death. Accordingly, for a model with death without preceding appropriate ICD therapy outcome, sustained VT/VF with appropriate ICD therapy event was censored at the arrhythmic event date. Alive and event-free patients were censored at the date of the last device follow-up.

We compared the strength of association of demographic, clinical, ECG, and device characteristics between the two competing outcomes (appropriate ICD therapies and all-cause death) by testing the null hypothesis that the coefficients for a given variable (*b*_*1*_ and *b*_*2*_) were the same across the two competing outcomes by calculating Z-scores:

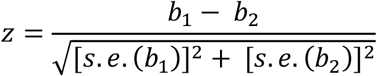

where *s*.*e*. is a standard error, and determining their statistical significance.

To study an association of GEH with the primary competing outcomes, we constructed four models. All models (1-4) with GEH exposure variables were adjusted for the type of median beat, mean RR interval, presence of PVCs, and possible distortion of median beat by premature beats. Model 1 was adjusted for demographic characteristics (age, sex, race) and study center. Model 2 included model 1 variables with additional adjustment for known clinical risk factors of the primary competing outcomes [LVEF, NYHA class, CM type, hypertension, diabetes, stroke, atrial fibrillation, eGFR_CKD-EPI_, and medications (use of beta-blockers, ACEi/ARBs, and class 1 or III antiarrhythmic drugs (AAD)]. Model 3 contained all covariates included in model 2, and, in addition, device characteristics: device type (ICD or CRT-D), manufacturer, and programming (VT and VF zone cutoffs and detection duration category). Model 4, in addition to model 3 variables, was adjusted for traditional ECG metrics: heart rate, QRS duration, and Hodges-corrected QT interval. To standardize comparisons, we expressed continuous variables per 1 SD.

To investigate previously reported discrepancy in the association of SAIQRST with ventricular tachyarrhythmias/SCD between studies that included entirely or mostly infarct-related CM (ICM) patients^3, 24^ and studies that included mixed ICM and NICM patients,^16, 25^ we tested the interaction of CM type with SAIQRST and VMQTi.

To appropriately consider nonlinear (e.g., U-shaped) hazards, associations of continuous GEH variables with primary competing outcomes were also studied using adjusted (model 1) Cox regression models incorporating cubic splines with 4 knots.

To test the study findings’ robustness under the missing at random assumption, we conducted a sensitivity analysis using an imputed dataset based on complete ECG data. The dataset without missing pre-implant ECG data included 2251 patients; 480 of them had sustained VT/VF with appropriate ICD therapy, and 401 died without appropriate ICD therapy.

Statistical analyses were performed using STATA MP 16.1 (StataCorp LP, College Station, TX). A *P* value < 0.05 was considered statistically significant. STATA do files are available at https://github.com/Tereshchenkolab/gehco.

## Results

### Study Population

After excluding the ineligible patient records and those with missing outcomes, our study population included 2,668 patients (Figure 2). Clinical characteristics are shown in Table 1.

**Table 1.** Clinical characteristics of the study population.

| Characteristic | N=2668 |
| --- | --- |
| Age(SD), y | 63.1(13.0) |
| Female, % | 23.4 |
| White, % | 78.4 |
| Black, % | 9.7 |
| Hispanic,% | 7.5 |
| Ischemic cardiomyopathy, % | 57.0 |
| Nonischemic cardiomyopathy, % | 43.0 |
| LVEF(SD), % | 28.2(11.1) |
| NYHA class I, % | 14.5 |
| NYHA class II, % | 39.6 |
| NYHA III, % | 43.2 |
| NYHA IV, % | 2.7 |
| Myocardial infarction, % | 49.1 |
| History of revascularization, % | 43.1 |
| Hypertension, % | 69.4 |
| Diabetes, % | 36.2 |
| Atrial fibrillation, % | 33.9 |
| History of stroke, % | 11.1 |
| Class I or III antiarrhythmic drugs, % | 11.2 |
| RAAS-modifying drugs, % | 78.7 |
| Use of beta-blockers, % | 85.1 |
| eGFR<sub>CKD-EPI</sub>(SD), mL/min/1.73 m<sup>2</sup> | 70.4(29.0) |
| BUN(SD), mg/dL | 24.7(13.8) |
| ICD single-chamber, % | 36.2 |
| ICD dual-chamber, % | 26.9 |
| CRT-D, % | 36.3 |
| ICD replacement, % | 30.2 |
| VT detection zone programmed, % | 90.5 |
| Antitachycardia pacing ON, % | 85.8 |
| VT zone (SD), bpm | 178(15) |
| VF zone (SD), bpm | 225.4(48.3) |
| Historic ICD therapies programming, % | 67.3 |
| Delayed ICD therapies programming, % | 32.7 |
| Medtronic device, % | 60.5 |
| Boston Scientific device, % | 24.3 |
| St. Jude device, % | 13.4 |

**Figure 2.**
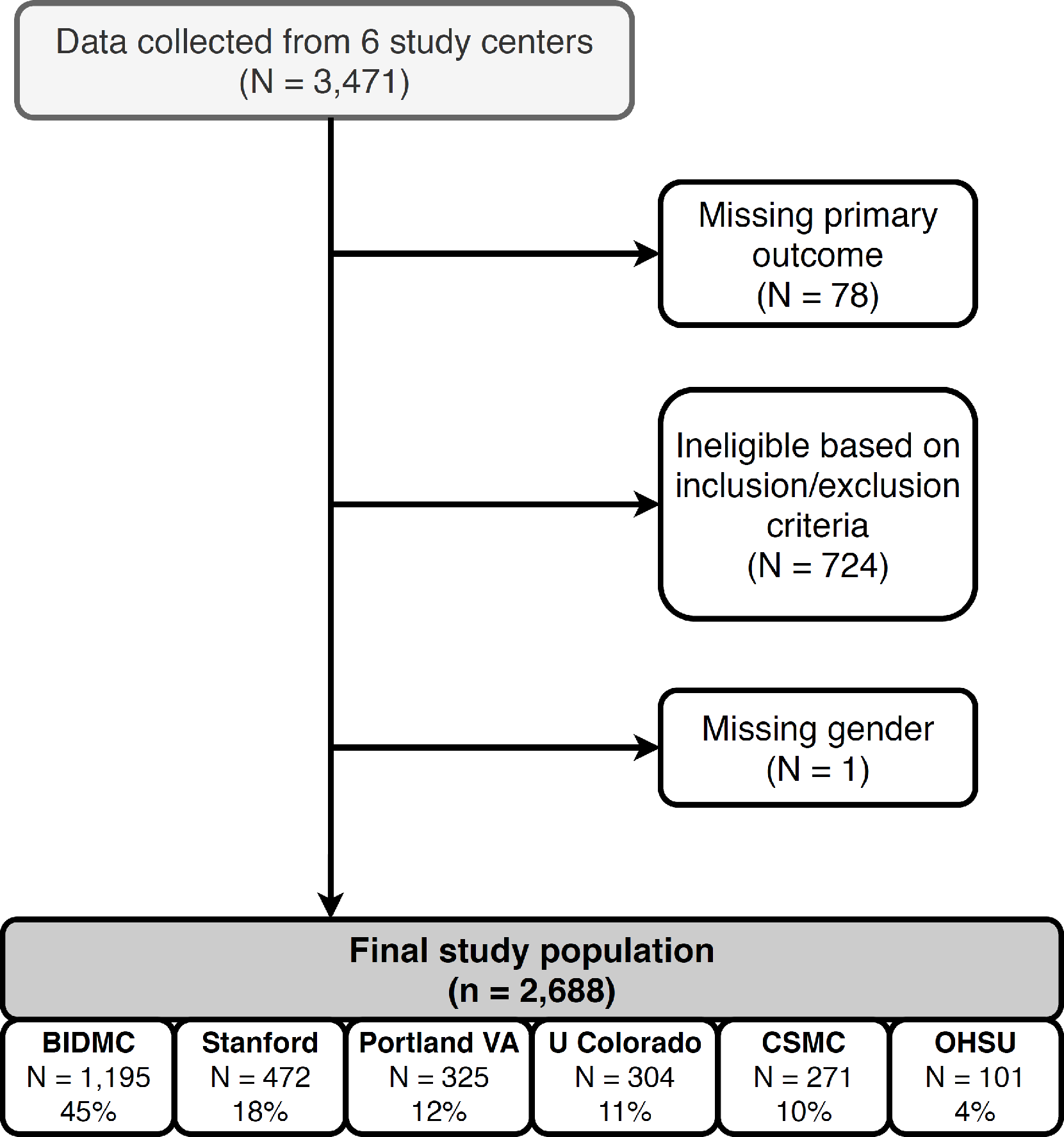
Study flowchart.

Over a median retrospective follow-up of 4 years, 541 patients experienced sustained ventricular tachyarrhythmia leading to appropriate ICD therapy [incidence rate 50.1 (95% CI 46.0-54.5) per 1000 person-years], and 479 patients died without appropriate ICD therapy [incidence rate 44.0 (95%CI 40.2-48.1) per 1000 person-years]. The average rate of appropriate ICD therapy was similar to the rate of death without appropriate ICD therapy: ∼ 5% per year.

An average heart rate, QRS duration, and QTc were within the normal range (Table 2). Patients who died without appropriate ICD therapy had a higher heart rate than those who remained alive and event-free (P=0.030). Patients who received appropriate ICD therapies were more likely to have PVC recorded on 12-lead ECG, smaller SVG magnitude and SAIQRST, wider QRS-T angle, and SVG vector pointing more superiorly, in comparison to events-free patients.

**Table 2.** Comparison of ECG and VCG metrics in patients with vs. without primary competing outcomes.

| Characteristic | Outcome-free<br>(n=1,351) | Appropriate VTVF<br>therapy (n=478) | Death before<br>VTVF (n=401) | P-value |
| --- | --- | --- | --- | --- |
| Heart Rate(SD), bpm | 71.0(14.8) | 71.5(15.7) | 73.2(16.2) | 0.037 |
| QRS duration(SD), ms | 116(35) | 118(38) | 120(35) | 0.178 |
| QTc Bazett(SD), ms | 463.7(51.3) | 452.6(52.4) | 465.0(51.6) | 0.0001 |
| QTc Fridericia(SD), ms | 452.2(47.8) | 441.1(49.7) | 451.4(49.6) | 0.0001 |
| QTc Hodges(SD), ms | 450.8(45.9) | 440.6(48.3) | 450.1(47.6) | 0.0002 |
| Median beat N, n(%) | 1034(76.5)) | 352(73.6) | 269(67.1) | 0.001 |
| Median beat S, n(%) | 154(11.4) | 71(14.9) | 61(15.2) |  |
| Median beat VP, n(%) | 163(12.1) | 55(11.5) | 71(17.7) |  |
| PVC presence, n(%) | 229(17.0) | 116(24.3) | 77(19.2) | 0.002 |
| SAIQRST(SD), mVms | 199.7(114.0) | 180.8(102.7) | 198.2(107.1) | 0.005 |
| VMQTi(SD), mVms | 136.1(79.7) | 121.7(69.0) | 133.4(71.6) | 0.002 |
| Area SVG (SD), mVms | 44.2(26.7) | 34.9(19.0) | 37.1(21.2) | <0.0001 |
| Peak SVG magnitude(SD), mV | 1.19(0.50) | 1.12(0.44) | 1.09(0.46) | 0.0001 |
| Area QRS-T angle(95%CI), deg | 142.7(140.9-144.4) | 146.9(144.5-149.4) | 149.0(146.5-151.5) | 0.005 |
| Peak QRS-T angle(95%CI), deg | 135.4(133.0-137.8) | 139.5(136.1-142.8) | 143.6(140.3-146.9) | <0.0001 |
| Area SVG elevation(95%CI),° | 85.6(84.0-87.2) | 90.2(87.2-93.3) | 90.2(87.3-93.1) | 0.147 |
| Peak SVG elevation(95%CI),° | 82.5.(81.3-83.8) | 83.9(81.5-86.2) | 86.9(84.3-89.4) | 0.029 |
| Area SVG azimuth(95%CI),° | 52.9(49.4-56.5) | 57.7(50.8-64.7) | 70.7(63.6-77.8) | 0.001 |
| Peak SVG azimuth(95%CI),° | 60.6(58.2-63.1) | 60.2(55.9-64.6) | 64.2(59.5-68.8) | 0.318 |
| T area(SD), mVms | 47.1(34.6) | 42.6(31.1) | 48.9(32.7) | 0.011 |
| Peak T magnitude(SD), mV | 0.34(0.22) | 0.32(0.21) | 0.35(0.21) | 0.074 |
| QRS area(SD), mVms | 72.1(50.2) | 61.0(40.0) | 67.9(42.1) | 0.0001 |
| Peak QRS magnitude(SD), mV | 1.41(0.57) | 1.33(0.51) | 1.33(0.53) | 0.005 |

Median beat: N=normal sinus; S=atrial fibrillation or flutter; VP=ventricular paced

### Demographic and clinical characteristics associated with primary competing outcomes

In minimally adjusted model 1 (Supplemental Table 3), male sex, black race, atrial fibrillation, use of AADs, history of MI and revascularization procedures, NYHA class III, and higher BUN were associated with increased risk of appropriate ICD therapies. In contrast, greater age, eGFR, and LVEF were associated with a lower risk of appropriate ICD therapies. Adjustment for cardiac disease substrate, risk factors (model 2), and device programming (model 3) explained the association of race, atrial fibrillation, MI and revascularization, NYHA class, and BUN with sustained VT/VF, which became non-significant. In fully adjusted analysis (model 4), only age, male sex, use of AADs, LVEF, and eGFR remained significantly associated with sustained VT/VF and appropriate ICD therapies (Supplemental Table 3).

In model 1 assessing the competing outcome of death without appropriate ICD therapy (Supplemental Table 4), age, ICM, MI, revascularization history, NYHA class II-IV, diabetes, atrial fibrillation, and BUN were associated with a higher risk of all-cause death, while use of beta-blockers, ACEIs, higher LVEF, and higher eGFR were associated with a lower risk of death. Adjustment for the HF characteristics and risk factors (model 2), device characteristics (model 3), and traditional ECG metrics (model 4) only slightly attenuated an association of clinical risk factors with the competing mortality outcome.

Even after comprehensive adjustment (model 3), several clinical risk factors had significantly different cause-specific hazards associated with the two competing outcomes (Supplemental Table 5). For every 13 years (1 SD) increase in age, the hazard of appropriate ICD therapy decreased by 20% (Figure 3), whereas the hazard of death without appropriate ICD therapy increased by 32%. Diabetes increased the hazard of death without appropriate ICD therapy by 45% but was not associated with appropriate ICD therapy. As expected, NYHA class and BUN had stronger associations with death than with appropriate ICD therapy. The use of beta-blockers and ACEIs was associated with a reduced hazard of death but not with the hazard of appropriate ICD therapy.

**Figure 3.**
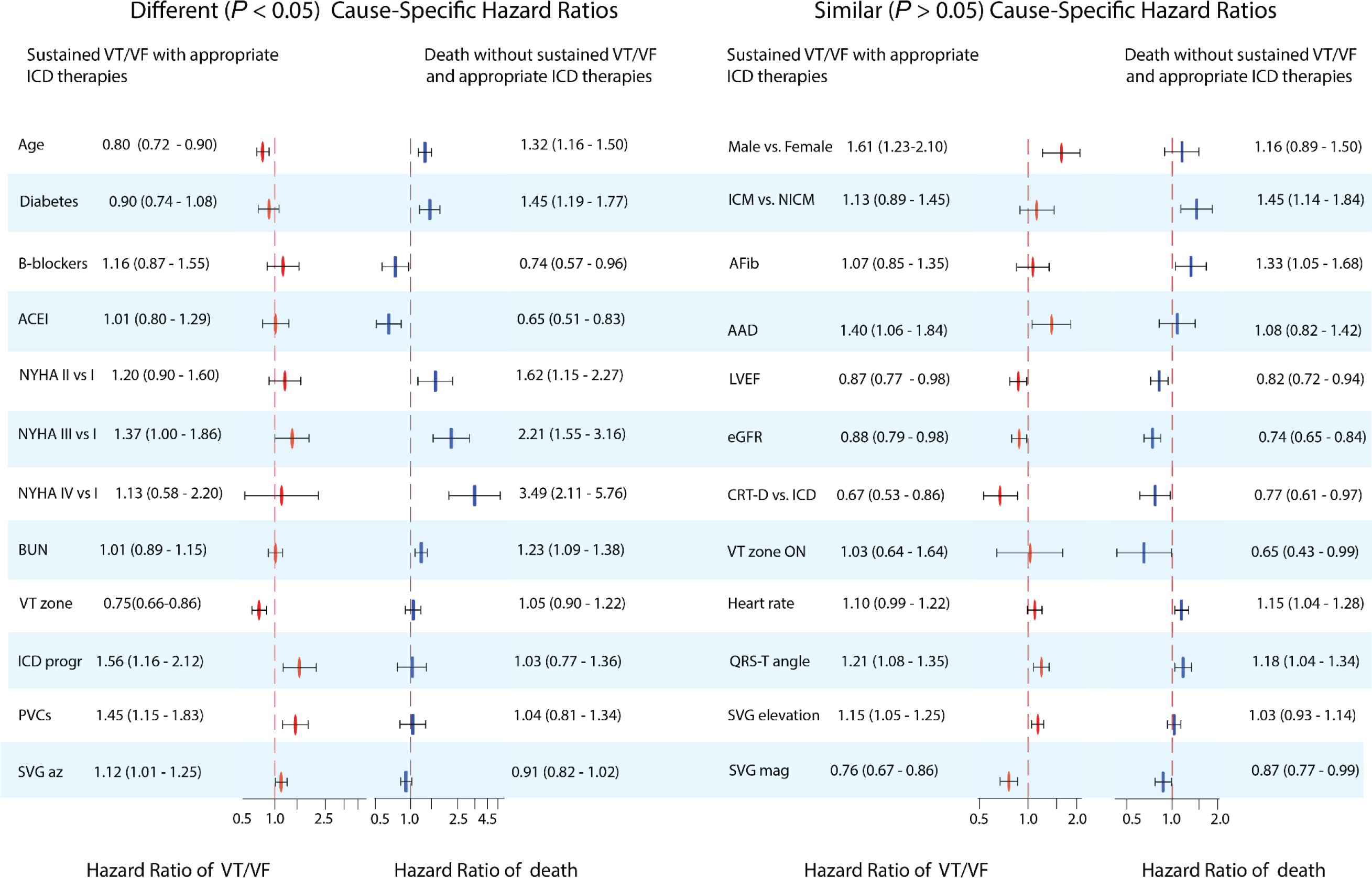
Statistically significantly different (*P*<0.05) or similar (*P*≥0.05) adjusted (model 3) cause-specific Cox hazard ratios and 95% confidence interval of two competing outcomes: sustained ventricular tachyarrhythmia with appropriate ICD therapies (red ovals) and all-cause death without prior ventricular tachyarrhythmia with appropriate ICD therapies (blue rectangles).

In contrast, there was no statistically significant difference in adjusted cause-specific hazards of two competing outcomes for male sex, ischemic CM, atrial fibrillation, the use of AADs, LVEF, and eGFR (Figure 3).

### Device characteristics associated with primary competing outcomes

In minimally adjusted model 1, ICD device type and programming were significantly associated with sustained VT/VF and appropriate ICD therapies (Supplemental Table 3). Adjustment for HF characteristics, risk factors, comorbidities, and traditional ECG metrics (models 2-4) did not change the strength of the association. In contrast, very few device characteristics were associated with mortality without appropriate ICD therapy (Supplemental Table 4).

In adjusted competing risk analysis, device characteristics had significantly different cause-specific hazards associated with the two competing outcomes (Supplemental Table 5). Unsurprisingly, for each additional 15 bpm increase in VT zone threshold, the hazard of sustained ventricular tachyarrhythmias with appropriate ICD therapies decreased by 25% (Figure 3), but it did not affect all-cause mortality without sustained VT/VF. Consistent with prior observations, more aggressive ICD programming was associated with a 56% higher risk of appropriate ICD therapies but was not associated with the competing death outcome.

CRT-D implantation was associated with a lower risk of both competing outcomes (Figure 3).

### Association of GEH with primary competing outcomes

In minimally adjusted analysis, the presence of PVCs and all GEH metrics were associated with sustained VT/VF and appropriate ICD therapies (Supplemental Table 3). However, the adjustment for known clinical risk factors and device characteristics explained that association for SAIQRST and VMQTi. Notably, after full adjustment in model 4 (including heart rate, QTc, and QRS duration), area and peak spatial QRS-T angle and SVG elevation, area SVG magnitude, and peak SVG azimuth were associated with appropriate ICD therapies.

In comparison, only resting heart rate, spatial QRS-T angle, and area SVG magnitude associated with the competing mortality outcome. The strength of associations diminished from model 1 to model 4, suggesting confounding by the HF substrate (Supplemental Table 4).

In minimally adjusted competing risk analysis, heart rate and QTc had significantly different cause-specific hazards associated with the two competing outcomes (Supplemental Table 5). However, these differences were explained by differences in clinical risk factors. In adjusted models, QRS duration and corrected QT interval were not associated with either appropriate ICD therapies or total mortality. After adjustment, only PVCs and peak SVG azimuth had significantly different cause-specific hazards associated with two competing outcomes (Figure 3). The presence of PVCs was associated with a 42% higher risk of appropriate ICD therapies but did not associate with competing mortality risk.

Each additional 1 SD increase in peak SVG azimuth (+36°, backward) was associated with a 12% increase in the hazard of appropriate ICD therapies and a 10% reduction in the hazard of total mortality without appropriate ICD therapy. Accordingly, 1 SD reduction in peak SVG azimuth (−36°, forward) was associated with a 12% decrease in the hazard of appropriate ICD therapies and a 10% increase in the risk of death without appropriate ICD therapy. Figure 4 illustrates opposing directions of hazards of two competing outcomes across peak SVG azimuth distribution. Area SVG azimuth showed a U-shaped association with appropriate ICD therapy (Supplemental Table 3) but not competing death risk (Supplemental Table 4).

**Figure 4.**
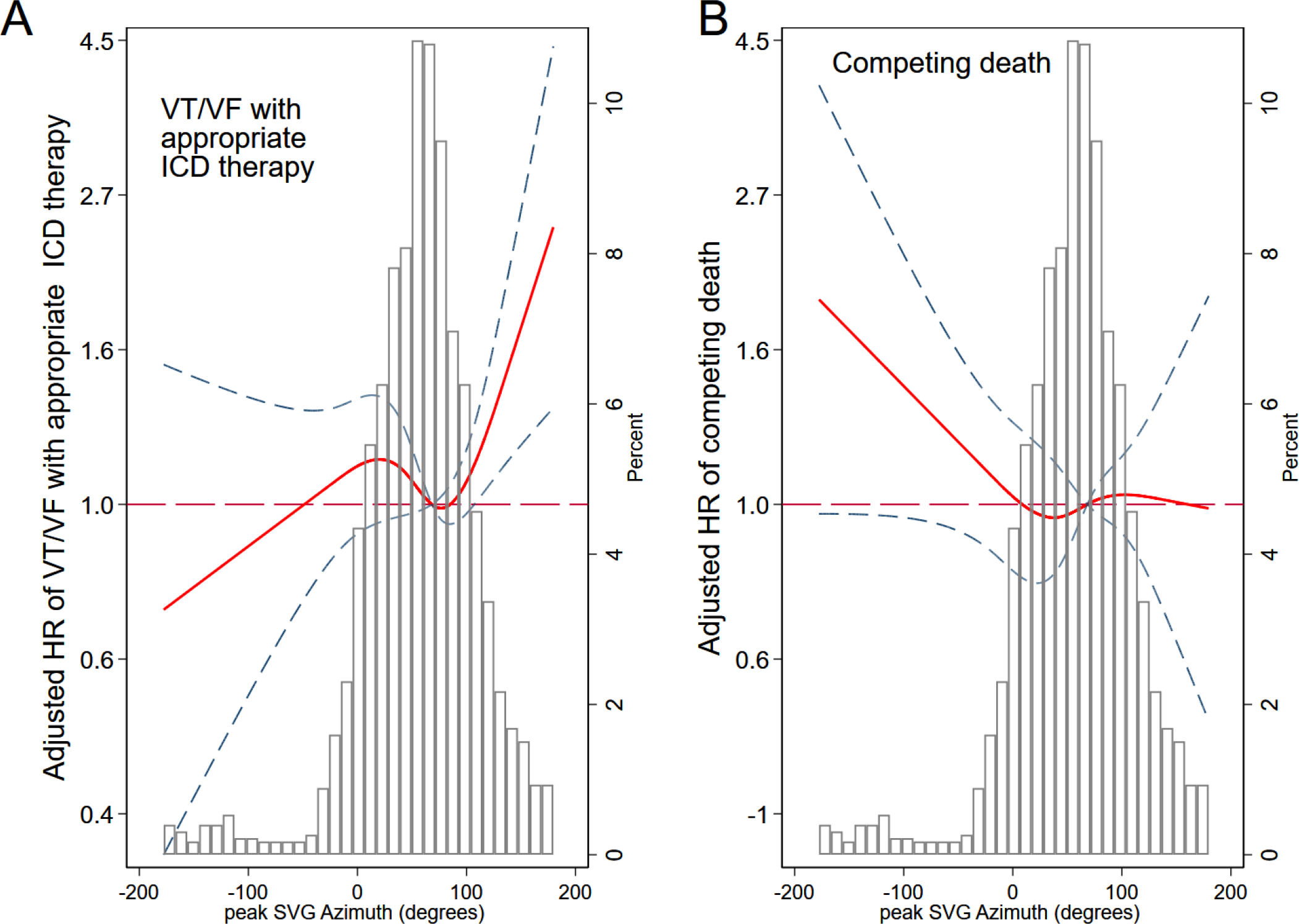
Adjusted (model 1) risk of (**A**) sustained VT/VF with appropriate ICD therapy and (**B**) competing death without appropriate ICD therapy associated with peak SVG azimuth. Restricted cubic spline with 95% confidence interval shows a change in cause-specific Cox hazard ratio (HR) (Y-axis) in response to peak SVG azimuth change (X-axis). The 50^th^ percentile of the SVG azimuth is selected as a reference. Knots of the peak SVG azimuth are at (−21)-42-75-137 degrees.

Spatial QRS-T angle and SVG magnitude were associated with both appropriate ICD therapy and death without appropriate ICD therapy. For each 1 SD (33°) increase in area QRS-T angle, we observed an ∼20% increase in the hazard of both outcomes (Figure 3). Each additional 1 SD increase in area SVG magnitude (25 mV*ms) was associated with a 25% reduction in the hazard of appropriate ICD therapies and a 14% reduction in the hazard of death without appropriate ICD therapy. For area SVG elevation, each 1 SD (31°) increase was associated with a 14% increase in the hazard of appropriate ICD therapies.

Sensitivity analysis (Supplemental Table 6) showed similar results.

### Interaction of SAIQRST with cardiomyopathy type

Cardiomyopathy type significantly modified the association of SAIQRST (and VMQTi) with the primary arrhythmic (but not competing death) outcome (Supplemental Table 7). After adjustment for HF characteristics, risk factors, and comorbidities (model 3), for each additional 111 mVms increase in SAIQRST, the hazard of sustained VT/VF with appropriate ICD therapies *increased* by 29% in ICM patients, but *decreased* by 22% in NICM subgroup (Figure 5).

**Figure 5.**
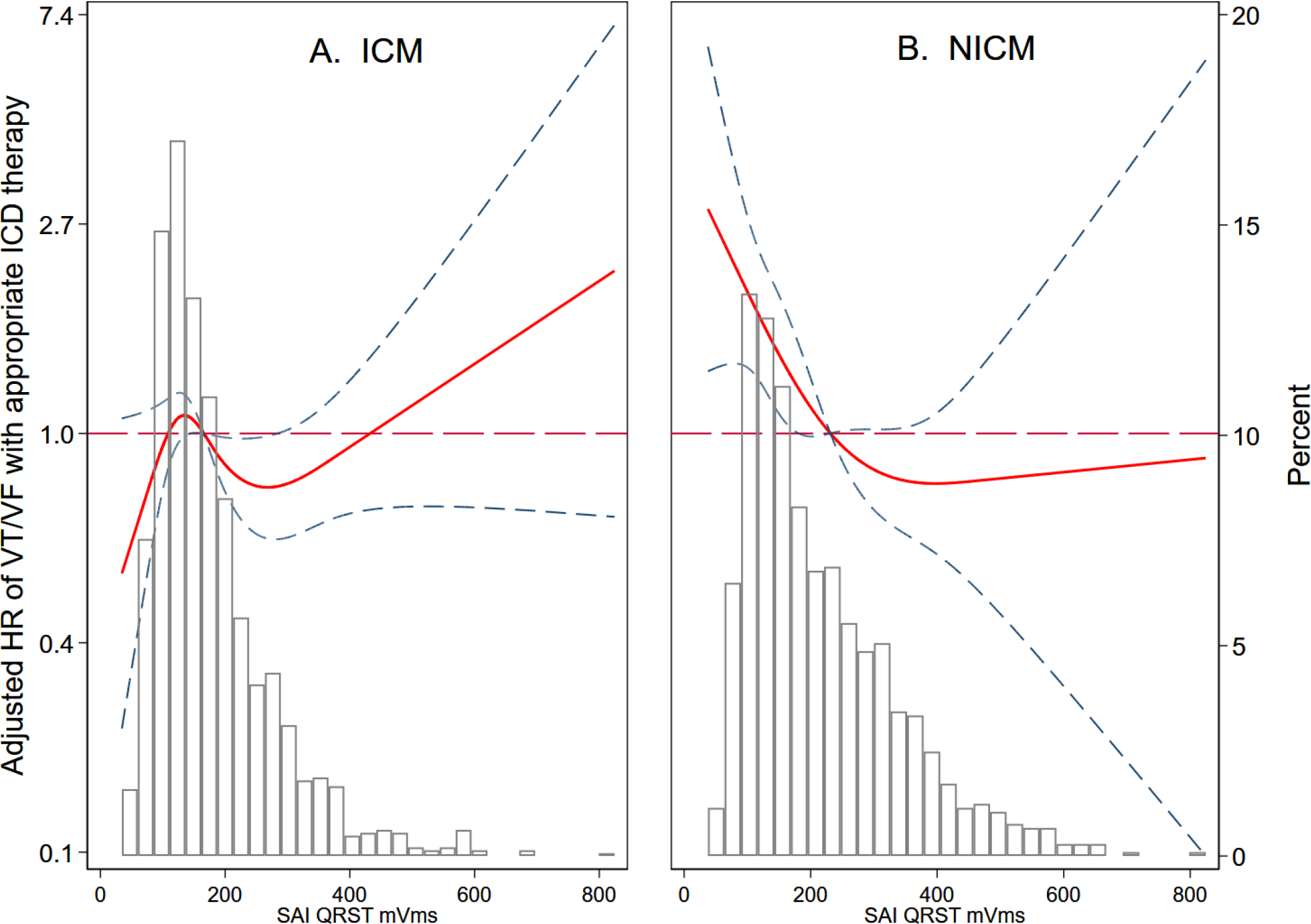
Adjusted (model 1) risk of sustained VT/VF with appropriate ICD therapy associated with sum absolute QRST integral (SAIQRST) in ischemic cardiomyopathy (**A**; ICM) and nonischemic cardiomyopathy (**B**; NICM). Restricted cubic spline with 95% confidence interval shows a change in cause-specific Cox hazard ratio (HR) (Y-axis) in response to SAIQRST change (X-axis). The 50^th^ percentile of the SAIQRST is selected as a reference. Knots of the SAIQRST in ICM patients are at 74-127-186-372 mVms, and in NICM patients are at 80-146-242-462 mVms.

## Discussion

This large multicenter retrospective cohort study of novel risk factors for SCD had two important findings. First, in competing risk analysis, we showed an independent association of GEH with ventricular arrhythmias leading to appropriate ICD therapies. After adjusting for demographic, clinical, ECG, and device characteristics, the SVG vector pointed in distinctly different directions in patients who developed sustained VT/VF rescued by appropriate ICD therapies (backward-upward, counterclockwise) versus those who died without ICD therapy (forward, clockwise). SVG magnitude and spatial QRS-T angle were associated with both competing outcomes. An association of SAIQRST with sustained VT/VF was modified by CM type. Smaller SAIQRST was associated with *increased* risk of VT/VF with appropriate ICD therapies in NICM, but *decreased* risk in ICM.

Second, we showed significantly different strength of association of age, diabetes, use of beta-blockers and ACEIs, NYHA class, BUN, ICD device programming features and VT zone cutoff, and the presence of PVCs between the two competing outcomes (appropriate ICD therapies and all-cause death). There was no difference in the associations of sex, CM type, atrial fibrillation, AAD use, LVEF, eGFR, and ICD device type with both competing outcomes.

### Differences in the risk factors of two competing outcomes

The importance of considering naturally competing outcomes in ICD patients is critically important.^26^ However, very few studies conducted a formal comparison of the strengths of association of a risk factor with two competing outcomes.^27^ In our study of primary prevention ICD recipients, unsurprisingly, VT/VF detection duration and VT zone cutoff were distinctly different attributes of the risk of ventricular tachyarrhythmia with appropriate ICD therapies.^28^ After adjustment for HF characteristics, comorbidities, and management, each additional 15 bpm increase in VT zone threshold was associated with reducing the hazard of sustained ventricular tachyarrhythmias and appropriate ICD therapies by 25%. Long detection duration reduced the hazard of appropriate ICD therapy by 58%. Our results are consistent with the well-recognized fact that appropriate ICD therapies occur more frequently than SCD,^29^ and ICD programming determines whether patients receive unnecessary therapies for what would have been nonsustained VT with longer ICD detection times.^28^

We observed that diabetes, BUN, NYHA class, and HF therapy (ACEIs, beta-blockers) were strongly associated with all-cause death without appropriate ICD therapies but *did not* associate with the primary arrhythmic outcome. These clinical characteristics are often perceived as risk factors of both outcomes. Observed differences in these clinical risk factors’ association with two competing outcomes should be considered in planning future clinical studies and developing SCD risk models.

Age was associated with both outcomes in distinctly different ways. After adjusting for confounders, for every 1 SD (13 years) increase in age, the hazard of death without preceding appropriate ICD therapy *increased* by 35%, while the hazard of sustained ventricular tachyarrhythmia leading to appropriate ICD therapy *decreased* by 21%. Age is a ubiquitous risk marker. It is well-recognized that many ICD recipients, despite having an ICD, often die secondary to progressive heart failure or other non-cardiac comorbidities that congregate with advanced age.

### Similarities in the risk factors of two competing outcomes

Noticeably, a set of particular clinical risk factors (sex, type of CM, atrial fibrillation, use of AADs, LVEF, eGFR), and ICD type had similar associations with both outcomes, suggesting common or overlapping mechanisms. All of these risk factors are well-known; however, our study conducted a novel formal statistical comparison of the strengths of their associations with competing outcomes in primary prevention ICD recipients. Common mechanisms of two competing outcomes highlight challenges in selecting an ideal candidate for primary prevention ICD.

### Global electrical heterogeneity

Our study confirmed that after multivariable adjustment in competing risk analysis, there is an independent association between SVG direction and sustained ventricular tachyarrhythmias leading to appropriate ICD therapies, but not the competing death. The SVG vector points towards the area of the myocardium with the shortest excited state,^4, 10^ and deviations from normal suggest the accumulation of a critical mass of abnormal electrical substrate, which might predispose to ventricular arrhythmias. Our results support the use of GEH, and specifically the orientation of the area SVG vector, as a marker of abnormal underlying electrophysiological substrate responsible for a propensity for sustained ventricular arrhythmias.^8, 9^ Further studies of mechanisms behind SVG directions are warranted.

Notably, we observed that the presence of PVCs on the 12-lead ECG was also associated with the competing risk of appropriate ICD therapy, but not all-cause mortality. Cardiac memory developing in response to PVCs may affect GEH.^30^ It is well known that PVCs are associated with the development or worsening of HF, and that PVCs can trigger life-threatening ventricular tachyarrhythmias. Further studies of the interaction between an underlying arrhythmogenic substrate (manifesting by GEH) and PVCs (manifesting by cardiac memory development) are needed to uncover the underlying mechanisms, leading to novel therapies.

In contrast to SVG direction, which had significantly different cause-specific hazards associated with two competing outcomes, area SVG magnitude and spatial QRS-T angle had similar associations with both outcomes. The finding of QRS-T angle being associated with both competing outcomes is expected. Numerous previous studies have shown an association between QRS-T angle and broadly defined cardiovascular disease.^31^ Our results underscore the importance and added value of a comprehensive GEH concept, which includes measurement of SVG direction, in addition to the QRS-T angle.

Our results also explain a previously observed discrepancy in SAIQRST. Similar to this study, in another primary prevention ICD study that enrolled NICM patients (PROSE-ICD),^16, 25^ smaller SAIQRST was associated with increased risk of appropriate ICD therapies. In contrast, in the ICM MADIT-II study,^24^ and the general population,^3^ *larger* SAIQRST was associated with ventricular arrhythmias and SCD. The current study demonstrated an opposite association of SAIQRST with sustained VT/VF and appropriate ICD therapies in patients with ICM or NICM.

In our previous studies of healthy individuals^11^ and the general population,^7, 22, 23^ we primarily observed agreement between findings obtained by using area-based and peak-based GEH metrics. However, in this study of HF patients with reduced LVEF, there were some differences in the results conveyed by peak-based and area-based SVG azimuth and magnitude and QRS-T angle. Theoretically, such differences can be explained by the underlying biophysical and physiological meaning. By measuring peak QRS and peak T vectors, we assess the moment when most of the heart is depolarized (QRS) or repolarized (T).^32, 33^ By measuring QRS and T areas, we aim to assess the complete depolarization (QRS) and repolarization (T) phase.

However, in the diseased/failing hearts, we may not be able to accurately identify an onset and offset of depolarization and repolarization in the entire heart by measuring the onset and offset of QRS and T on the surface ECG due to the heterogeneity and multipolar nature of these processes.^34^ A single-diploe ECG approximation carries inherent limitation in the modeling of multipolar electrical activity in diseased hearts.^34^ In the diseased hearts, peak-based measurements can be more meaningful and carry less error than area-based GEH metrics.

### Statistical assessment of competing risks

In this study, we preferred to use cause-specific hazards rather than the analysis of subdistribution hazards because Fine and Gray’s method is not valid for causal inference and can produce misleading results.^35, 36^ We used cause-specific hazard that removes an individual from the risk set when *any* type of event occurs. In contrast, the subdistribution hazard does not remove an individual from the risk set when a competing event occurs. Nonetheless, those who experience one competing event are no longer at risk of the other competing event. As a result, the Fine and Gray method does not isolate distinct causal effects on the competing risks but instead may confound them. The interpretation of Fine and Gray’s method is challenging when outcomes are not rare (as in this study).^37^ On the other hand, cumulative incidence functions are useful for prediction, which we utilized in our previous study where we developed a GEH-based SCD competing risk.^3^

### Strengths and Limitations

Our study’s strengths include the large sample size and population drawn from a diverse group of medical centers from across the United States. However, because the study included only academic medical centers, the results may not apply to all patients with ICDs. Because of our study’s retrospective nature, ICD programming was not standardized, and differences in ICD detection and treatment parameters could have influenced the results, although we attempted to correct this by including ICD programming in our multivariable models. There was no postmortem ICD interrogation data, and we cannot precisely determine if there were ventricular arrhythmias close to the time of death. Importantly, appropriate ICD therapy is a surrogate forSCD, as all ventricular arrhythmias treated by the ICD would not necessarily have led to cardiac arrest or SCD. As in any observational study, residual confounding cannot be completely eliminated.

## Data Availability

The data will be available through the National Heart, Lung, and Blood Institute Biological Specimen and Data Repository Information Coordinating Center (BioLINCC) after the publication of this manuscript (planning submission). The open-source MATLAB (MathWorks, Natick, MA, USA) code for ECG analysis is provided at https://physionet.org/physiotools/geh and https://github.com/Tereshchenkolab/Origin. Statistical analysis code is provided at https://github.com/Tereshchenkolab/statistics.

https://github.com/Tereshchenkolab/Origin

https://github.com/Tereshchenkolab/statistics

## Abbreviations

CM: cardiomyopathy
LVEF: left ventricular ejection fraction
SCD: sudden cardiac death
ICD: implantable cardioverter-defibrillator
NYHA: New York Heart Association
HF: heart failure
ICM: ischemic/infarct-related cardiomyopathy
NICM: nonischemic cardiomyopathy
GEH: global electrical heterogeneity
SVG: spatial ventricular gradient
CRT-D: cardiac resynchronization therapy defibrillator
PVC: premature ventricular complex
SAIQRST: sum absolute QRST integral
VMQTi: QT integral on vector magnitude signal
NID: number of intervals to detect

## Acknowledgment

The authors thank Zachary Zouyed, BS, Nichole Rogovoy, BS, Katherine Yang, BS, and Christopher Hamilton, BA, for their help with the data preparation for analysis.

## SUPPLEMENTAL MATREIAL

**Supplemental Table 1.** ICD VT/VF detection duration programming categories.

| Possible programming options (either/or) | Long detection duration | Short detection duration |
| --- | --- | --- |
| Initial NID VT, n | $\geq 30$ | $< 30$ |
| Redetect NID VT, n | $\geq 18$ | $< 18$ |
| NID numerator, n | $\geq 30$ | $< 30$ |
| NID denominator, n | $\geq 40$ | $< 40$ |
| Initial time VT, sec | $\geq 10$ | $< 10$ |
| Redetect time VT, sec | $> 1$ | $\leq 1$ |
| Initial time VF, sec | $\geq 3$ | $< 3$ |
| Redetect time VF, sec | $> 1$ | $\leq 1$ |

NID=number of intervals to detect.

**Supplemental Table 2.**
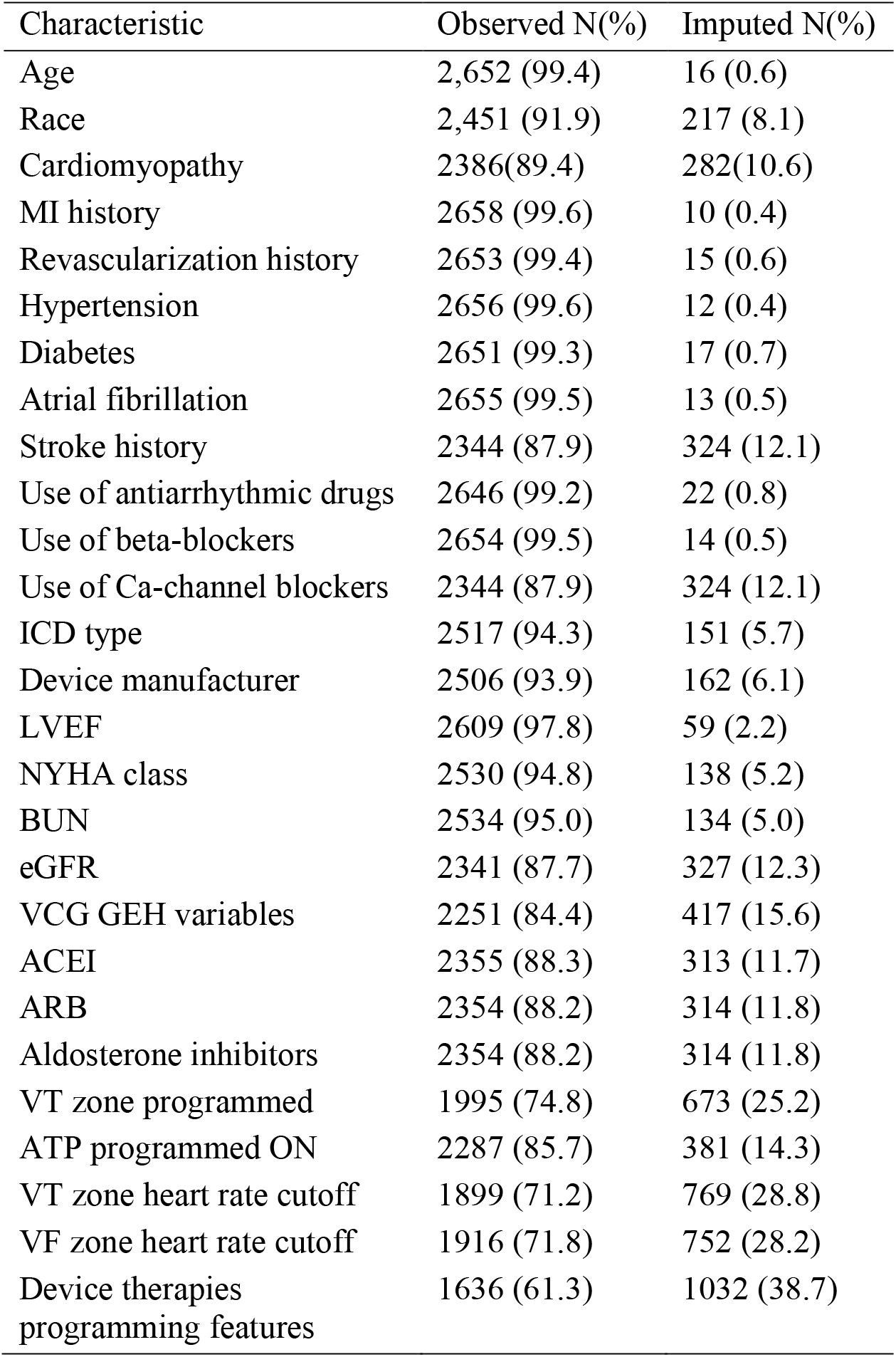
Summary of observed and imputed data.

| Characteristic | Observed N(%) | Imputed N(%) |
| --- | --- | --- |
| Age | 2,652 (99.4) | 16 (0.6) |
| Race | 2,451 (91.9) | 217 (8.1) |
| Cardiomyopathy | 2386(89.4) | 282(10.6) |
| MI history | 2658 (99.6) | 10 (0.4) |
| Revascularization history | 2653 (99.4) | 15 (0.6) |
| Hypertension | 2656 (99.6) | 12 (0.4) |
| Diabetes | 2651 (99.3) | 17 (0.7) |
| Atrial fibrillation | 2655 (99.5) | 13 (0.5) |
| Stroke history | 2344 (87.9) | 324 (12.1) |
| Use of antiarrhythmic drugs | 2646 (99.2) | 22 (0.8) |
| Use of beta-blockers | 2654 (99.5) | 14 (0.5) |
| Use of Ca-channel blockers | 2344 (87.9) | 324 (12.1) |
| ICD type | 2517 (94.3) | 151 (5.7) |
| Device manufacturer | 2506 (93.9) | 162 (6.1) |
| LVEF | 2609 (97.8) | 59 (2.2) |
| NYHA class | 2530 (94.8) | 138 (5.2) |
| BUN | 2534 (95.0) | 134 (5.0) |
| eGFR | 2341 (87.7) | 327 (12.3) |
| VCG GEH variables | 2251 (84.4) | 417 (15.6) |
| ACEI | 2355 (88.3) | 313 (11.7) |
| ARB | 2354 (88.2) | 314 (11.8) |
| Aldosterone inhibitors | 2354 (88.2) | 314 (11.8) |
| VT zone programmed | 1995 (74.8) | 673 (25.2) |
| ATP programmed ON | 2287 (85.7) | 381 (14.3) |
| VT zone heart rate cutoff | 1899 (71.2) | 769 (28.8) |
| VF zone heart rate cutoff | 1916 (71.8) | 752 (28.2) |
| Device therapies<br>programming features | 1636 (61.3) | 1032 (38.7) |

**Supplemental Table 3.** The cause-specific hazard of sustained ventricular tachyarrhythmia with appropriate ICD therapy.

| Predictor | Model 1 |  | Model 2 |  | Model 3 |  | Model 4 |  |
| --- | --- | --- | --- | --- | --- | --- | --- | --- |
|  | HR (95%CI) | P | HR (95%CI) | P | HR (95%CI) | P | HR (95%CI) | P |
| **Age, per 1SD (13y)** | **0.88(0.80-0.97)** | **0.010** | **0.80(0.71-0.89)** | **<0.0001** | **0.80(0.72-0.90)** | **<0.0001** | **0.79(0.70-0.89)** | **<0.0001** |
| **Male sex (vs. female)** | **1.63(1.26-2.11)** | **<0.0001** | **1.59(1.22-2.07)** | **0.001** | **1.61(1.23-2.10)** | **0.001** | **1.55(1.19-2.03)** | **0.001** |
| Black race (vs. white) | **1.45(1.08-1.95)** | **0.014** | **1.40(1.03-1.91)** | **0.030** | 1.37(0.99-1.87) | 0.054 | 1.31(0.95-1.81) | 0.096 |
| **ICM (vs. NICM)** | 1.24(0.99-1.56) | 0.061 | **1.30(1.02-1.64)** | **0.031** | 1.13(0.89-1.45) | 0.318 | 1.17(0.91-1.51) | 0.220 |
| Hypertension | 1.01(0.83-1.24) | 0.890 | 0.97(0.79-1.20) | 0.804 | 0.96(0.78-1.19) | 0.696 | 0.96(0.78-1.19) | 0.705 |
| **Diabetes** | **0.99(0.82-1.18)** | **0.892** | **0.91(0.75-1.10)** | **0.323** | **0.90(0.74-1.08)** | **0.256** | **0.89(0.73-1.08)** | **0.230** |
| Atrial fibrillation | **1.25(1.01-1.54)** | **0.041** | 1.08(0.86-1.37) | 0.487 | 1.07(0.85-1.35) | 0.583 | 1.09(0.86-1.38) | 0.460 |
| Stroke | 1.20(0.92-1.56) | 0.187 | 1.22(0.93-1.61) | 0.146 | 1.18(0.90-1.56) | 0.228 | 1.21(0.92-1.60) | 0.176 |
| **AAD (class I or III) use** | **1.56(1.23-2.00)** | **<0.0001** | **1.40(1.07-1.83)** | **0.015** | **1.40(1.06-1.84)** | **0.016** | **1.52(1.15-2.01)** | **0.003** |
| **Beta-blockers use** | **1.19(0.90-1.56)** | **0.228** | **1.15(0.86-1.52)** | **0.340** | **1.16(0.87-1.55)** | **0.303** | **1.16(0.87-1.55)** | **0.304** |
| **LVEF, per 1SD (11%)** | **0.83(0.74-0.92)** | **<0.0001** | **0.86(0.77-0.97)** | **0.012** | **0.87(0.77-0.98)** | **0.020** | **0.89(0.79-0.997)** | **0.045** |
| NYHA class II vs. I | 1.21(0.92-1.60) | 0.165 | 1.14(0.86-1.51) | 0.375 | 1.20(0.90-1.60) | 0.220 | 1.21(0.90-1.61) | 0.203 |
| **NYHA class III vs. I** | **1.36(1.03-1.79)** | **0.030** | **1.17(0.87-1.58)** | **0.291** | **1.37(1.00-1.86)** | **0.048** | **1.34(0.98-1.84)** | **0.066** |
| **NYHA class IV vs. I** | **1.28(0.68-2.41)** | **0.453** | **1.07(0.56-2.05)** | **0.846** | **1.13(0.58-2.20)** | **0.716** | **0.97(0.49-1.90)** | **0.924** |
| **eGFR, per 1SD (29 mL/min/1.73 m²)** | **0.84(0.75-0.93)** | **0.001** | **0.87(0.78-0.97)** | **0.016** | **0.88(0.79-0.98)** | **0.023** | **0.88(0.78-0.98)** | **0.020** |
| **ACE Inhibitors** | **1.12(0.92-1.36)** | **0.254** | **1.00(0.79-1.26)** | **0.981** | **1.01(0.80-1.29)** | **0.909** | **1.03(0.81-1.32)** | **0.784** |
| Angiotensin Receptor Blockers use | 0.82(0.65-1.06) | 0.129 | 0.84(0.62-1.12) | 0.234 | 0.91(0.68-1.23) | 0.549 | 0.93(0.69-1.26) | 0.640 |
| **CRT-D vs. ICD** | **0.77(0.61-0.95)** | **0.017** | **0.68(0.53-0.86)** | **0.002** | **0.67(0.53-0.86)** | **0.002** | **0.68(0.52-0.89)** | **0.005** |
| **VT zone, per 1SD (15 bpm)** | **0.75(0.67-0.85)** | **<0.0001** | **0.77(0.68-0.87)** | **<0.0001** | **0.75(0.66-0.86)** | **<0.0001** | **0.75(0.65-0.86)** | **<0.0001** |
| VF zone, per 1SD (48 bpm) | **0.88(0.77-1.004)** | **0.057** | 0.89(0.78-1.03) | 0.110 | 1.07(0.91-1.26) | 0.403 | 1.07(0.90-1.25) | 0.449 |
| **ICD therapy historic programming** | **1.59(1.20-2.10)** | **0.001** | **1.51(1.13-2.01)** | **0.005** | **1.56(1.16-2.12)** | **0.004** | **1.58(1.17-2.14)** | **0.003** |
| Heart rate, per 1SD (15 bpm) | **1.10(0.999-1.21)** | **0.051** | 1.09(0.99-1.21) | 0.083 | 1.10(0.99-1.22) | 0.075 | 1.09(0.98-1.20) | 0.125 |
| QTc Hodges, per 1SD (46 ms) | **0.92(0.84-1.02)** | **0.104** | **0.89(0.80-0.98)** | **0.024** | 0.93(0.83-1.03) | 0.148 | 0.90(0.81-1.01) | 0.077 |
| **PVC on 12-lead ECG** | **1.34(1.08-1.68)** | **0.009** | **1.38(1.10-1.72)** | **0.005** | **1.45(1.15-1.83)** | **0.002** | **1.42(1.13-1.79)** | **0.003** |
| QRS duration, per 1SD(36 ms) | 0.98(0.90-1.06) | 0.646 | 0.97(0.89-1.06) | 0.533 | 1.02(0.93-1.12) | 0.649 | 1.05(0.96-1.16) | 0.297 |
| **Myocardial infarction** | **1.34(1.11-1.61)** | **0.003** | 1.28(0.95-1.74) | 0.108 | 1.20(0.88-1.64) | 0.239 | 1.20(0.87-1.65) | 0.263 |
| **Revascularization** | **1.28(1.06-1.55)** | **0.009** | 1.21(0.94-1.56) | 0.140 | 1.23(0.95-1.59) | 0.112 | 1.24(0.96-1.61) | 0.106 |
| Ca-channel blockers use | 0.76(0.53-1.08) | 0.129 | 0.79(0.55-1.13) | 0.198 | 0.81(0.56-1.16) | 0.248 | 0.82(0.57-1.18) | 0.284 |
| Aldosterone antagonists use | 1.13(0.93-1.37) | 0.224 | 1.04(0.85-1.27) | 0.726 | 1.07(0.87-1.32) | 0.512 | 1.05(0.85-1.29) | 0.666 |
| **BUN, per 1SD (14 mg/dL)** | **1.11(1.02-1.22)** | **0.016** | **1.00(0.88-1.13)** | **0.985** | **1.01(0.89-1.15)** | **0.853** | **1.01(0.89-1.15)** | **0.850** |
| ATP programmed ON | 0.93(0.69-1.26) | 0.637 | 0.95(0.69-1.29) | 0.730 | 1.07(0.78-1.51) | 0.620 | 1.06(0.76-1.49) | 0.713 |
| VT zone programmed ON | 0.94(0.67-1.32) | 0.716 | 0.95(0.67-1.34) | 0.766 | 1.03(0.64-1.64) | 0.908 | 1.05(0.65-1.68) | 0.846 |
| **Area QRS-T angle, per 1SD (33°)** | **1.18(1.06-1.31)** | **0.002** | **1.17(1.05-1.31)** | **0.004** | **1.21(1.08-1.35)** | **0.001** | **1.21(1.08-1.36)** | **0.001** |
| **Peak QRS-T angle, per 1SD(43°)** | **1.11(1.01-1.22)** | **0.034** | 1.10(0.995-1.22) | 0.062 | **1.14(1.03-1.26)** | **0.015** | **1.13(1.02-1.26)** | **0.023** |
| **SAIQRST, per 1SD (111 mVms)** | **0.89(0.81-0.99)** | **0.025** | 0.90(0.81-1.004) | 0.058 | 0.95(0.85-1.06) | 0.374 | 0.92(0.80-1.06) | 0.248 |
| **VMQTi, per 1SD (80 mVms)** | **0.88(0.80-0.98)** | **0.016** | **0.89(0.80-0.99)** | **0.036** | 0.94(0.84-1.05) | 0.292 | 0.91(0.79-1.04) | 0.176 |
| **Peak SVG azimuth, per 1SD (36°)** | **1.12(1.01-1.24)** | **0.033** | **1.09(0.98-1.20)** | **0.108** | **1.12(1.01-1.25)** | **0.031** | **1.12(1.01-1.24)** | **0.039** |
| Area SVG azimuth T1(-180-20) | 1.19(0.95-1.50) | 0.132 | 1.24(0.98-1.56) | 0.080 | 1.15(0.90-1.47) | 0.262 | 1.16(0.91-1.49) | 0.234 |
| Area SVG azimuth T2(20-70) | Reference |  | Reference |  | Reference |  | Reference |  |
| Area SVG azimuth T3(70-180) | 1.25(0.99-1.58) | 0.056 | 1.21(0.96-1.52) | 0.115 | 1.20(0.95-1.52) | 0.134 | 1.20(0.95-1.52) | 0.128 |
| Area SVG azimuth Q1 (-180-0) | 1.21(0.93-1.57) | 0.152 | 1.22(0.93-1.59) | 0.155 | 1.14(0.87-1.50) | 0.349 | 1.14(0.87-1.50) | 0.339 |
| Area SVG azimuth Q2 (0-50) | Reference |  | Reference |  | Reference |  | Reference |  |
| Area SVG azimuth Q3 (50-90) | 1.12(0.86-1.45) | 0.397 | 1.07(0.82-1.39) | 0.635 | 1.11(0.84-1.45) | 0.462 | 1.09(0.83-1.43) | 0.549 |
| Area SVG azimuth Q4 (90-180) | 1.19(0.91-1.55) | 0.211 | 1.10(0.84-1.45) | 0.497 | 1.11(0.85-1.47) | 0.439 | 1.10(0.84-1.46) | 0.480 |
| **Area SVG elevation, per 1SD (31°)** | **1.13(1.03-1.24)** | **0.007** | **1.13(1.03-1.23)** | **0.010** | **1.15(1.05-1.25)** | **0.003** | **1.14(1.04-1.25)** | **0.004** |
| **Peak SVG elevation, per 1SD (25°)** | 1.10(0.999-1.21) | 0.053 | 1.10(0.99-1.20) | 0.079 | **1.10(1.004-1.21)** | **0.042** | **1.11(1.005-1.22)** | **0.040** |
| **Area SVG magnitude, per 1SD (25mVms)** | **0.72(0.64-0.81)** | **<0.0001** | **0.73(0.65-0.83)** | **<0.0001** | **0.76(0.67-0.86)** | **<0.0001** | **0.75(0.66-0.85)** | **<0.0001** |
| Peak SVG magnitude, per 1SD (0.5mV) | 0.92(0.83-1.2) | 0.097 | 0.95(0.86-1.05) | 0.336 | 0.95(0.86-1.06) | 0.381 | 0.95(0.86-1.06) | 0.377 |

Red color indicates a statistically significant difference in the point estimates for two competing outcomes. Yellow and green colors
indicate models with SVG azimuth tertiles (T1-T2-T3) and quartiles (Q1-Q2-Q3-Q4). Gray color indicates variables included in each model.

**Supplemental Table 4.** The cause-specific hazard of all-cause death without appropriate ICD therapy.

| Predictor | Model 1 |  | Model 2 |  | Model 3 |  | Model 4 |  |
| --- | --- | --- | --- | --- | --- | --- | --- | --- |
|  | HR (95%CI) | P | HR (95%CI) | P | HR (95%CI) | P | HR (95%CI) | P |
| **Age, per 1SD (13y)** | **1.61(1.44-1.80)** | **<0.0001** | **1.29(1.14-1.47)** | **<0.0001** | **1.32(1.16-1.50)** | **<0.0001** | **1.35(1.18-1.55)** | **<0.0001** |
| Male sex (vs. female) | 1.17(0.91-1.49) | 0.228 | 1.17(0.90-1.52) | 0.241 | 1.16(0.89-1.50) | 0.280 | 1.16(0.89-1.51) | 0.280 |
| Black race (vs. white) | 1.13(0.78-1.62) | 0.516 | 1.03(0.70-1.51) | 0.884 | 0.99(0.67-1.45) | 0.945 | 0.95(0.64-1.41) | 0.809 |
| **ICM (vs. NICM)** | **1.43(1.14-1.79)** | **0.002** | **1.50(1.19-1.90)** | **0.001** | **1.45(1.14-1.84)** | **0.002** | **1.43(1.12-1.82)** | **0.004** |
| Hypertension | 1.14(0.91-1.43) | 0.250 | 0.95(0.74-1.20) | 0.652 | 0.97(0.76-1.24) | 0.809 | 0.98(0.77-1.25) | 0.881 |
| **Diabetes** | **1.81(1.51-2.18)** | **<0.0001** | **1.47(1.21-1.80)** | **<0.0001** | **1.45(1.19-1.77)** | **<0.0001** | **1.41(1.15-1.72)** | **0.001** |
| **Atrial fibrillation** | **1.48(1.20-1.84)** | **<0.0001** | **1.33(1.06-1.69)** | **0.015** | **1.33(1.05-1.68)** | **0.019** | **1.35(1.07-1.71)** | **0.012** |
| Stroke | 1.22(0.91-1.62) | 0.179 | 1.19(0.88-1.59) | 0.257 | 1.17(0.87-1.57) | 0.298 | 1.19(0.88-1.59) | 0.257 |
| **AAD (class I, III) use** | **1.44(1.13-1.85)** | **0.004** | 1.06(0.80-1.39) | 0.693 | 1.08(0.82-1.42) | 0.602 | 1.10(0.83-1.45) | 0.512 |
| **Beta-blockers use** | **0.72(0.57-0.92)** | **0.008** | **0.74(0.58-0.95)** | **0.019** | **0.74(0.57-0.96)** | **0.021** | **0.75(0.58-0.96)** | **0.022** |
| **LVEF, per 1SD (11%)** | **0.74(0.66-0.84)** | **<0.0001** | **0.84(0.73-0.96)** | **0.009** | **0.82(0.72-0.94)** | **0.004** | **0.84(0.73-0.96)** | **0.013** |
| **NYHA class II vs. I** | **1.62(1.16-2.26)** | **0.004** | **1.54(1.10-2.17)** | **0.012** | **1.62(1.15-2.27)** | **0.006** | **1.61(1.14-1.27)** | **0.007** |
| **NYHA class III vs. I** | **2.51(1.82-3.47)** | **<0.0001** | **1.99(1.42-2.81)** | **<0.0001** | **2.21(1.55-3.16)** | **<0.0001** | **2.13(1.49-3.05)** | **<0.0001** |
| **NYHA class IV vs. I** | **4.35(2.71-6.98)** | **<0.0001** | **3.19(1.94-5.24)** | **<0.0001** | **3.49(2.11-5.76)** | **<0.0001** | **3.01(1.80-5.05)** | **<0.0001** |
| **eGFR, per 1SD (29 mL/min/1.73 m<sup>2</sup>)** | **0.65(0.57-0.73)** | **<0.0001** | **0.75(0.66-0.85)** | **<0.0001** | **0.74(0.65-0.84)** | **<0.0001** | **0.74(0.65-0.84)** | **<0.0001** |
| **ACE Inhibitors use** | **0.70(0.57-0.86)** | **0.001** | **0.66(0.52-0.84)** | **0.001** | **0.65(0.51-0.83)** | **0.001** | **0.68(0.53-0.88)** | **0.003** |
| Angiotensin receptor blockers use | 0.94(0.73-1.21) | 0.644 | 0.79(0.58-1.08) | 0.142 | 0.79(0.58-1.08) | 0.143 | 0.81(0.59-1.11) | 0.190 |
| **CRT-D vs. ICD** | 1.02(0.83-1.26) | 0.863 | **0.77(0.61-0.97)** | **0.029** | **0.77(0.61-0.97)** | **0.029** | 0.77(0.59-1.001) | 0.051 |
| **VT zone, per 1SD (15 bpm)** | **1.02(0.90-1.16)** | **0.759** | **1.07(0.94-1.21)** | **0.294** | **1.05(0.90-1.22)** | **0.545** | **1.04(0.90-1.21)** | **0.595** |
| **VF zone, per 1SD (48 bpm)** | **1.08(0.94-1.23)** | **0.303** | 1.06(0.92-1.22) | 0.384 | 1.05(0.88-1.24) | 0.609 | 1.05(0.89-1.25) | 0.545 |
| **ICD therapy historic programming** | **1.05(0.80-1.36)** | **0.738** | **1.00(0.76-1.31)** | **0.995** | **1.03(0.77-1.36)** | **0.849** | **1.03(0.78-1.37)** | **0.824** |
| **Heart rate, per 1SD (15 bpm)** | **1.31(1.18-1.44)** | **<0.0001** | **1.15(1.04-1.28)** | **0.008** | **1.15(1.04-1.28)** | **0.008** | **1.15(1.03-1.28)** | **0.010** |
| **QTc Hodges, per 1SD (46 ms)** | **1.07(0.96-1.18)** | **0.206** | 0.98(0.88-1.09) | 0.746 | 1.01(0.90-1.12) | 0.898 | 1.01(0.90-1.13) | 0.918 |
| **PVC on 12-lead ECG** | **0.99(0.78-1.26)** | **0.961** | **1.04(0.81-1.34)** | **0.740** | **1.04(0.81-1.34)** | **0.752** | **1.05(0.81-1.35)** | **0.737** |
| **QRS duration, per 1SD(36 ms)** | **0.99(0.90-1.08)** | **0.760** | 0.95(0.86-1.05) | 0.348 | 0.99(0.89-1.10) | 0.828 | 0.99(0.88-1.11) | 0.824 |
| **Myocardial infarction** | **1.28(1.04-1.56)** | **0.018** | 0.96(0.71-1.31) | 0.809 | 0.97(0.71-1.32) | 0.833 | 0.97(0.71-1.33) | 0.863 |
| **Revascularization** | **1.25(1.02-1.52)** | **0.029** | 0.95(0.74-1.22) | 0.660 | 0.94(0.73-1.21) | 0.620 | 0.95(0.74-1.23) | 0.718 |
| Ca-channel blockers use | 1.05(0.75-1.47) | 0.796 | 1.06(0.75-1.49) | 0.736 | 1.08(0.76-1.52) | 0.681 | 1.09(0.77-1.55) | 0.615 |
| Aldosterone antagonists use | 0.94(0.75-1.19) | 0.628 | **0.78(0.61-0.99)** | **0.038** | 0.79(0.62-1.001) | 0.051 | 0.79(0.62-1.01) | 0.061 |
| **BUN, per 1SD (14 mg/dL)** | **1.43(1.32-1.54)** | **<0.0001** | **1.21(1.08-1.36)** | **0.001** | **1.23(1.09-1.38)** | **0.001** | **1.24(1.10-1.39)** | **<0.0001** |
| ATP programmed ON | 0.78(0.60-1.01) | 0.057 | 0.81(0.62-1.06) | 0.117 | 0.78(0.59-1.03) | 0.078 | 0.79(0.59-1.05) | 0.098 |
| **VT zone programmed ON** | 0.79(0.58-1.06) | 0.119 | 0.79(0.58-1.08) | 0.133 | **0.65(0.43-0.99)** | **0.042** | 0.67(0.44-1.01) | 0.054 |
| **Area QRS-T angle, per 1SD (33°)** | **1.17(1.04-1.31)** | **0.007** | **1.15(1.01-1.29)** | **0.030** | **1.18(1.04-1.34)** | **0.011** | **1.20(1.05-1.37)** | **0.008** |
| **Peak QRS-T angle, per 1SD (43°)** | **1.16(1.04-1.30)** | **0.007** | **1.13(1.004-1.27)** | **0.042** | **1.16(1.03-1.31)** | **0.016** | **1.18(1.04-1.33)** | **0.012** |
| SAIQRST, per 1SD (111mVms) | 0.97(0.87-1.07) | 0.494 | 0.98(0.88-1.09) | 0.727 | 1.02(0.91-1.14) | 0.790 | 1.03(0.90-1.19) | 0.664 |
| VMQTi, per 1SD (80 mVms) | 0.96(0.87-1.06) | 0.416 | 0.97(0.87-1.09) | 0.639 | 1.01(0.90-1.13) | 0.866 | 1.02(0.89-1.18) | 0.752 |
| **Peak SVG azimuth, per 1SD (36°)** | **0.95(0.85-1.05)** | **0.293** | **0.90(0.81-1.0004)** | **0.051** | **0.91(0.82-1.02)** | **0.094** | **0.91(0.82-1.02)** | **0.095** |
| Area SVG azimuth T1(-180-20) | Reference |  | Reference |  | Reference |  | Reference |  |
| Area SVG azimuth T2(20-70) | 1.17(0.91-1.50) | 0.222 | 1.13(0.87-1.46) | 0.374 | 1.18(0.90-1.54) | 0.225 | 1.19(0.90-1.56) | 0.215 |
| Area SVG azimuth T3(70-180) | **1.32(1.03-1.70)** | **0.030** | 1.17(0.90-1.51) | 0.243 | 1.22(0.93-1.58) | 0.145 | 1.22(0.94-1.60) | 0.135 |
| Area SVG azimuth Q1 (-180-0) | Reference |  | Reference |  | Reference |  | Reference |  |
| Area SVG azimuth Q2 (0-50) | 1.08(0.81-1.45) | 0.601 | 1.07(0.79-1.45) | 0.676 | 1.11(0.81-1.52) | 0.506 | 1.11(0.81-1.52) | 0.505 |
| Area SVG azimuth Q3 (50-90) | 1.26(0.94-1.68) | 0.116 | 1.13(0.84-1.52) | 0.419 | 1.22(0.90-1.65) | 0.206 | 1.23(0.90-1.68) | 0.191 |
| **Area SVG azimuth Q4 (90-180)** | **1.39(1.04-1.85)** | **0.027** | 1.22(0.90-1.64) | 0.198 | 1.28(0.94-1.73) | 0.114 | 1.29(0.95-1.76) | 0.104 |
| Area SVG elevation, per 1SD (31°) | 1.03(0.94-1.14) | 0.499 | 1.02(0.92-1.13) | 0.743 | 1.03(0.93-1.14) | 0.564 | 1.03(0.93-1.15) | 0.524 |
| Peak SVG elevation, per 1SD (25°) | 1.06(0.95-1.17) | 0.315 | 1.01(0.91-1.13) | 0.803 | 1.02(0.92-1.14) | 0.712 | 1.02(0.92-1.14) | 0.686 |
| **Area SVG magnitude, per 1SD(25mVms)** | **0.81(0.72-0.91)** | **0.001** | **0.85(0.75-0.97)** | **0.012** | **0.87(0.77-0.99)** | **0.033** | **0.86(0.76-0.98)** | **0.029** |
| Peak SVG magnitude, per 1SD (0.5mV) | 0.91(0.81-1.03) | 0.131 | 1.00(0.89-1.13) | 0.967 | 1.00(0.89-1.13) | 0.983 | 1.00(0.89-1.13) | 0.987 |

Red color indicates a statistically significant difference in the point estimates for two competing outcomes. Yellow and green colors indicate models with SVG azimuth tertiles (T1-T2-T3) and quartiles (Q1-Q2-Q3-Q4). Gray color indicates variables included in each model.

**Supplemental Table 5.**
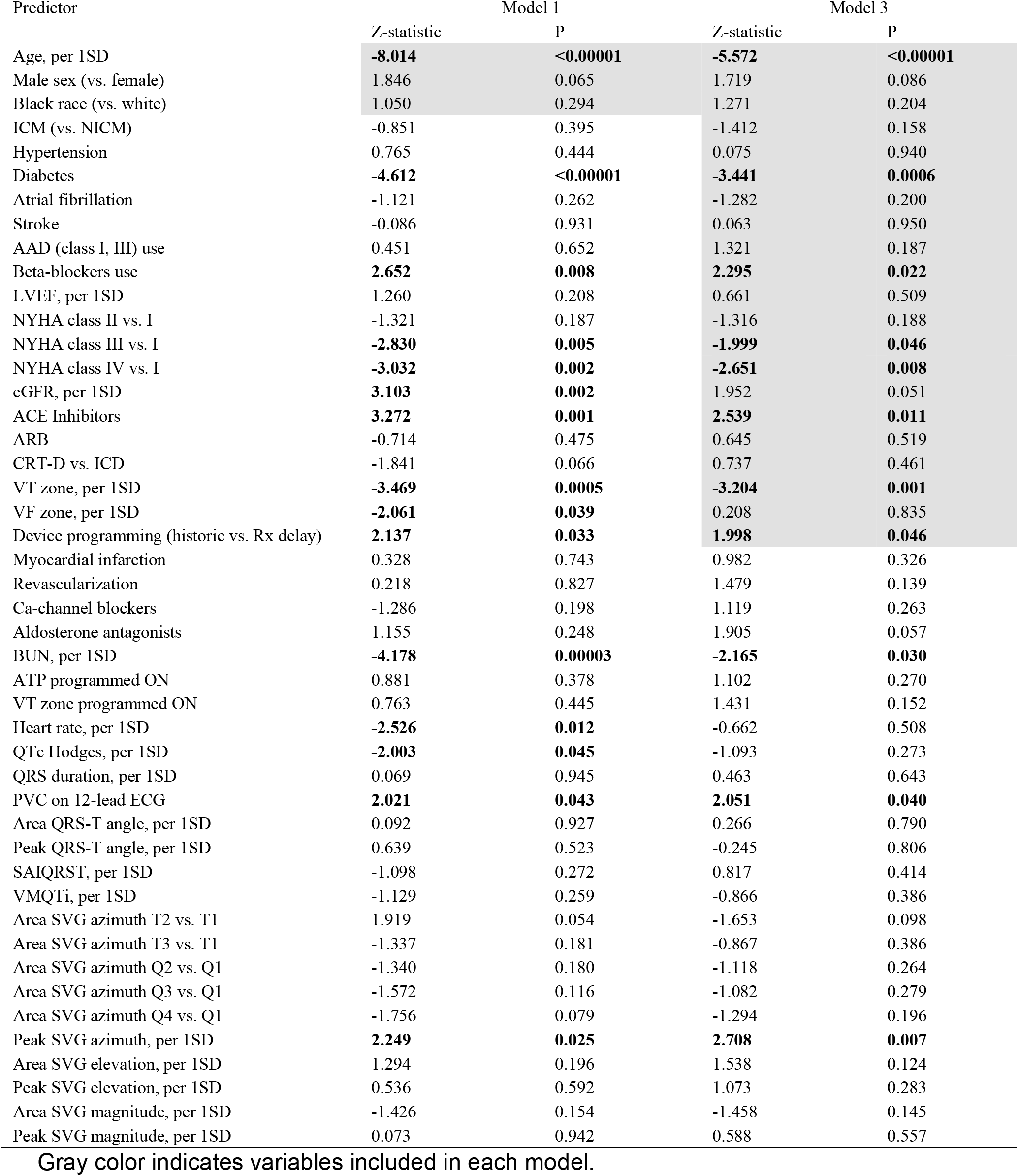
Statistics of Difference in the cause-specific hazards of two outcomes.

| Predictor | Model 1 |  | Model 3 |  |
| --- | --- | --- | --- | --- |
|  | Z-statistic | P | Z-statistic | P |
| Age, per 1SD | **-8.014** | **<0.00001** | **-5.572** | **<0.00001** |
| Male sex (vs. female) | 1.846 | 0.065 | 1.719 | 0.086 |
| Black race (vs. white) | 1.050 | 0.294 | 1.271 | 0.204 |
| ICM (vs. NICM) | -0.851 | 0.395 | -1.412 | 0.158 |
| Hypertension | 0.765 | 0.444 | 0.075 | 0.940 |
| Diabetes | **-4.612** | **<0.00001** | **-3.441** | **0.0006** |
| Atrial fibrillation | -1.121 | 0.262 | -1.282 | 0.200 |
| Stroke | -0.086 | 0.931 | 0.063 | 0.950 |
| AAD (class I, III) use | 0.451 | 0.652 | 1.321 | 0.187 |
| Beta-blockers use | **2.652** | **0.008** | **2.295** | **0.022** |
| LVEF, per 1SD | 1.260 | 0.208 | 0.661 | 0.509 |
| NYHA class II vs. I | -1.321 | 0.187 | -1.316 | 0.188 |
| NYHA class III vs. I | **-2.830** | **0.005** | **-1.999** | **0.046** |
| NYHA class IV vs. I | **-3.032** | **0.002** | **-2.651** | **0.008** |
| eGFR, per 1SD | **3.103** | **0.002** | 1.952 | 0.051 |
| ACE Inhibitors | **3.272** | **0.001** | **2.539** | **0.011** |
| ARB | -0.714 | 0.475 | 0.645 | 0.519 |
| CRT-D vs. ICD | -1.841 | 0.066 | 0.737 | 0.461 |
| VT zone, per 1SD | **-3.469** | **0.0005** | **-3.204** | **0.001** |
| VF zone, per 1SD | **-2.061** | **0.039** | 0.208 | 0.835 |
| Device programming (historic vs. Rx delay) | **2.137** | **0.033** | **1.998** | **0.046** |
| Myocardial infarction | 0.328 | 0.743 | 0.982 | 0.326 |
| Revascularization | 0.218 | 0.827 | 1.479 | 0.139 |
| Ca-channel blockers | -1.286 | 0.198 | 1.119 | 0.263 |
| Aldosterone antagonists | 1.155 | 0.248 | 1.905 | 0.057 |
| BUN, per 1SD | **-4.178** | **0.00003** | **-2.165** | **0.030** |
| ATP programmed ON | 0.881 | 0.378 | 1.102 | 0.270 |
| VT zone programmed ON | 0.763 | 0.445 | 1.431 | 0.152 |
| Heart rate, per 1SD | **-2.526** | **0.012** | -0.662 | 0.508 |
| QTc Hodges, per 1SD | **-2.003** | **0.045** | -1.093 | 0.273 |
| QRS duration, per 1SD | 0.069 | 0.945 | 0.463 | 0.643 |
| PVC on 12-lead ECG | **2.021** | **0.043** | **2.051** | **0.040** |
| Area QRS-T angle, per 1SD | 0.092 | 0.927 | 0.266 | 0.790 |
| Peak QRS-T angle, per 1SD | 0.639 | 0.523 | -0.245 | 0.806 |
| SAIQRST, per 1SD | -1.098 | 0.272 | 0.817 | 0.414 |
| VMQTi, per 1SD | -1.129 | 0.259 | -0.866 | 0.386 |
| Area SVG azimuth T2 vs. T1 | 1.919 | 0.054 | -1.653 | 0.098 |
| Area SVG azimuth T3 vs. T1 | -1.337 | 0.181 | -0.867 | 0.386 |
| Area SVG azimuth Q2 vs. Q1 | -1.340 | 0.180 | -1.118 | 0.264 |
| Area SVG azimuth Q3 vs. Q1 | -1.572 | 0.116 | -1.082 | 0.279 |
| Area SVG azimuth Q4 vs. Q1 | -1.756 | 0.079 | -1.294 | 0.196 |
| Peak SVG azimuth, per 1SD | **2.249** | **0.025** | **2.708** | **0.007** |
| Area SVG elevation, per 1SD | 1.294 | 0.196 | 1.538 | 0.124 |
| Peak SVG elevation, per 1SD | 0.536 | 0.592 | 1.073 | 0.283 |
| Area SVG magnitude, per 1SD | -1.426 | 0.154 | -1.458 | 0.145 |
| Peak SVG magnitude, per 1SD | 0.073 | 0.942 | 0.588 | 0.557 |

Gray color indicates variables included in each model.

**Supplemental Table 6.**
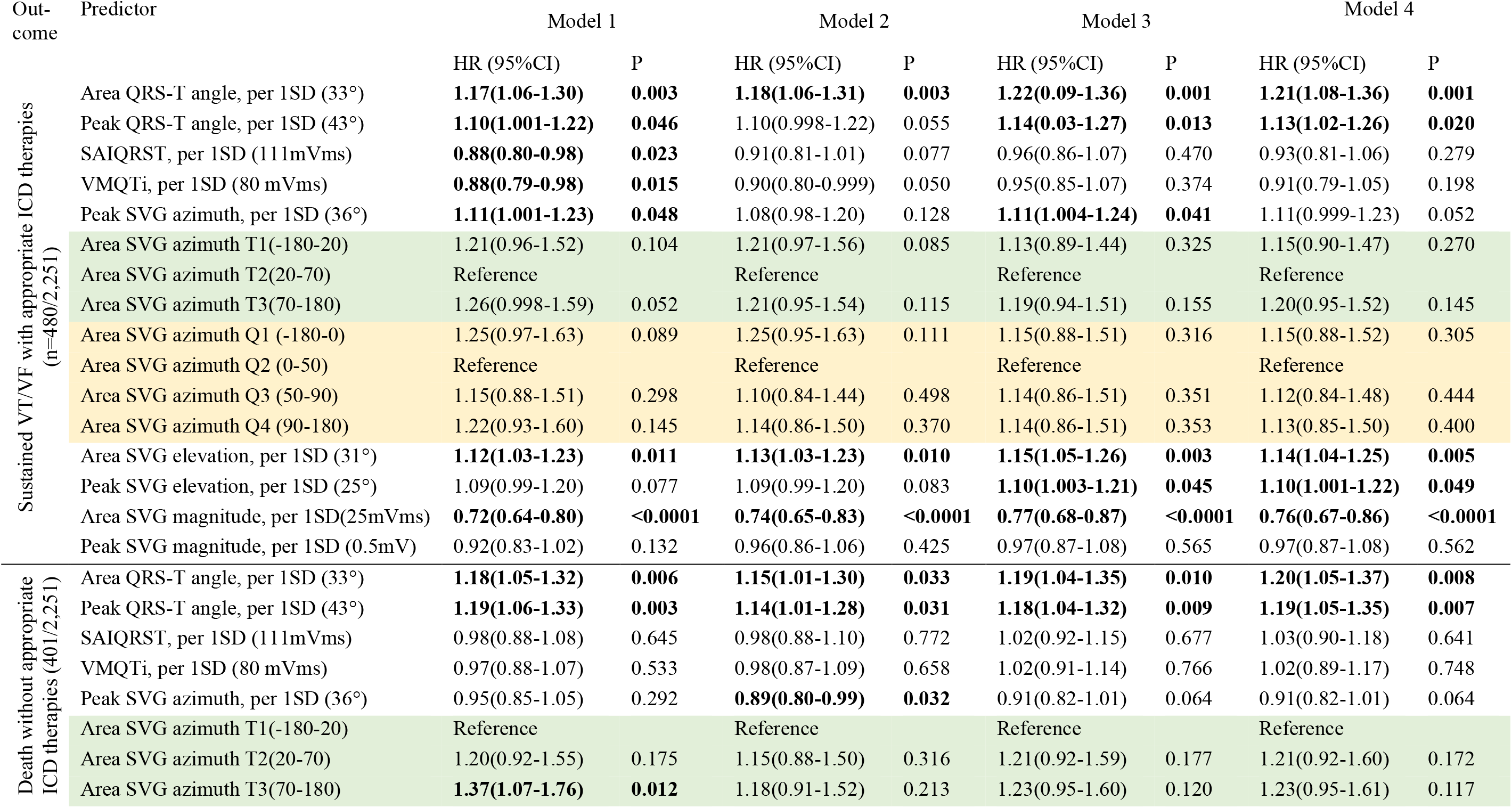
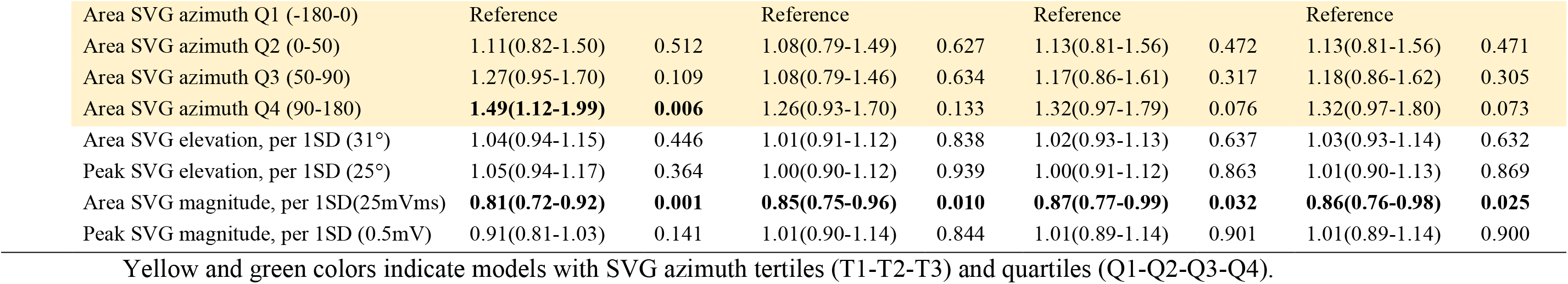
Sensitivity analysis using an imputed dataset based on complete ECG data (no ECG data were imputed): cause-specific hazard of competing outcomes.

| Out-<br>come | Predictor | Model 1 |  | Model 2 |  | Model 3 |  | Model 4 |  |
| --- | --- | --- | --- | --- | --- | --- | --- | --- | --- |
|  |  | HR (95%CI) | P | HR (95%CI) | P | HR (95%CI) | P | HR (95%CI) | P |
| Sustained VT/VF with appropriate ICD therapies<br>(n=480/2,251) | Area QRS-T angle, per 1SD (33°) | **1.17(1.06-1.30)** | **0.003** | **1.18(1.06-1.31)** | **0.003** | **1.22(0.09-1.36)** | **0.001** | **1.21(1.08-1.36)** | **0.001** |
|  | Peak QRS-T angle, per 1SD (43°) | **1.10(1.001-1.22)** | **0.046** | 1.10(0.998-1.22) | 0.055 | **1.14(0.03-1.27)** | **0.013** | **1.13(1.02-1.26)** | **0.020** |
|  | SAIQRST, per 1SD (111mVms) | **0.88(0.80-0.98)** | **0.023** | 0.91(0.81-1.01) | 0.077 | 0.96(0.86-1.07) | 0.470 | 0.93(0.81-1.06) | 0.279 |
|  | VMQTi, per 1SD (80 mVms) | **0.88(0.79-0.98)** | **0.015** | 0.90(0.80-0.999) | 0.050 | 0.95(0.85-1.07) | 0.374 | 0.91(0.79-1.05) | 0.198 |
|  | Peak SVG azimuth, per 1SD (36°) | **1.11(1.001-1.23)** | **0.048** | 1.08(0.98-1.20) | 0.128 | **1.11(1.004-1.24)** | **0.041** | 1.11(0.999-1.23) | 0.052 |
|  | Area SVG azimuth T1(-180-20) | 1.21(0.96-1.52) | 0.104 | 1.21(0.97-1.56) | 0.085 | 1.13(0.89-1.44) | 0.325 | 1.15(0.90-1.47) | 0.270 |
|  | Area SVG azimuth T2(20-70) | Reference |  | Reference |  | Reference |  | Reference |  |
|  | Area SVG azimuth T3(70-180) | 1.26(0.998-1.59) | 0.052 | 1.21(0.95-1.54) | 0.115 | 1.19(0.94-1.51) | 0.155 | 1.20(0.95-1.52) | 0.145 |
|  | Area SVG azimuth Q1 (-180-0) | 1.25(0.97-1.63) | 0.089 | 1.25(0.95-1.63) | 0.111 | 1.15(0.88-1.51) | 0.316 | 1.15(0.88-1.52) | 0.305 |
|  | Area SVG azimuth Q2 (0-50) | Reference |  | Reference |  | Reference |  | Reference |  |
|  | Area SVG azimuth Q3 (50-90) | 1.15(0.88-1.51) | 0.298 | 1.10(0.84-1.44) | 0.498 | 1.14(0.86-1.51) | 0.351 | 1.12(0.84-1.48) | 0.444 |
|  | Area SVG azimuth Q4 (90-180) | 1.22(0.93-1.60) | 0.145 | 1.14(0.86-1.50) | 0.370 | 1.14(0.86-1.51) | 0.353 | 1.13(0.85-1.50) | 0.400 |
|  | Area SVG elevation, per 1SD (31°) | **1.12(1.03-1.23)** | **0.011** | **1.13(1.03-1.23)** | **0.010** | **1.15(1.05-1.26)** | **0.003** | **1.14(1.04-1.25)** | **0.005** |
|  | Peak SVG elevation, per 1SD (25°) | 1.09(0.99-1.20) | 0.077 | 1.09(0.99-1.20) | 0.083 | **1.10(1.003-1.21)** | **0.045** | **1.10(1.001-1.22)** | **0.049** |
|  | Area SVG magnitude, per 1SD(25mVms) | **0.72(0.64-0.80)** | **<0.0001** | **0.74(0.65-0.83)** | **<0.0001** | **0.77(0.68-0.87)** | **<0.0001** | **0.76(0.67-0.86)** | **<0.0001** |
|  | Peak SVG magnitude, per 1SD (0.5mV) | 0.92(0.83-1.02) | 0.132 | 0.96(0.86-1.06) | 0.425 | 0.97(0.87-1.08) | 0.565 | 0.97(0.87-1.08) | 0.562 |
| Death without appropriate ICD therapies (401/2,251) | Area QRS-T angle, per 1SD (33°) | **1.18(1.05-1.32)** | **0.006** | **1.15(1.01-1.30)** | **0.033** | **1.19(1.04-1.35)** | **0.010** | **1.20(1.05-1.37)** | **0.008** |
|  | Peak QRS-T angle, per 1SD (43°) | **1.19(1.06-1.33)** | **0.003** | **1.14(1.01-1.28)** | **0.031** | **1.18(1.04-1.32)** | **0.009** | **1.19(1.05-1.35)** | **0.007** |
|  | SAIQRST, per 1SD (111mVms) | 0.98(0.88-1.08) | 0.645 | 0.98(0.88-1.10) | 0.772 | 1.02(0.92-1.15) | 0.677 | 1.03(0.90-1.18) | 0.641 |
|  | VMQTi, per 1SD (80 mVms) | 0.97(0.88-1.07) | 0.533 | 0.98(0.87-1.09) | 0.658 | 1.02(0.91-1.14) | 0.766 | 1.02(0.89-1.17) | 0.748 |
|  | Peak SVG azimuth, per 1SD (36°) | 0.95(0.85-1.05) | 0.292 | **0.89(0.80-0.99)** | **0.032** | 0.91(0.82-1.01) | 0.064 | 0.91(0.82-1.01) | 0.064 |
|  | Area SVG azimuth T1(-180-20) | Reference |  | Reference |  | Reference |  | Reference |  |
|  | Area SVG azimuth T2(20-70) | 1.20(0.92-1.55) | 0.175 | 1.15(0.88-1.50) | 0.316 | 1.21(0.92-1.59) | 0.177 | 1.21(0.92-1.60) | 0.172 |
|  | Area SVG azimuth T3(70-180) | **1.37(1.07-1.76)** | **0.012** | 1.18(0.91-1.52) | 0.213 | 1.23(0.95-1.60) | 0.120 | 1.23(0.95-1.61) | 0.117 |

|  |  |  |  |  |  |  |  |  |
| --- | --- | --- | --- | --- | --- | --- | --- | --- |
| Area SVG azimuth Q1 (-180-0) | Reference |  | Reference |  | Reference |  | Reference |  |
| Area SVG azimuth Q2 (0-50) | 1.11(0.82-1.50) | 0.512 | 1.08(0.79-1.49) | 0.627 | 1.13(0.81-1.56) | 0.472 | 1.13(0.81-1.56) | 0.471 |
| Area SVG azimuth Q3 (50-90) | 1.27(0.95-1.70) | 0.109 | 1.08(0.79-1.46) | 0.634 | 1.17(0.86-1.61) | 0.317 | 1.18(0.86-1.62) | 0.305 |
| Area SVG azimuth Q4 (90-180) | **1.49(1.12-1.99)** | **0.006** | 1.26(0.93-1.70) | 0.133 | 1.32(0.97-1.79) | 0.076 | 1.32(0.97-1.80) | 0.073 |
| Area SVG elevation, per 1SD (31°) | 1.04(0.94-1.15) | 0.446 | 1.01(0.91-1.12) | 0.838 | 1.02(0.93-1.13) | 0.637 | 1.03(0.93-1.14) | 0.632 |
| Peak SVG elevation, per 1SD (25°) | 1.05(0.94-1.17) | 0.364 | 1.00(0.90-1.12) | 0.939 | 1.00(0.91-1.12) | 0.863 | 1.01(0.90-1.13) | 0.869 |
| Area SVG magnitude, per 1SD(25mVms) | **0.81(0.72-0.92)** | **0.001** | **0.85(0.75-0.96)** | **0.010** | **0.87(0.77-0.99)** | **0.032** | **0.86(0.76-0.98)** | **0.025** |
| Peak SVG magnitude, per 1SD (0.5mV) | 0.91(0.81-1.03) | 0.141 | 1.01(0.90-1.14) | 0.844 | 1.01(0.89-1.14) | 0.901 | 1.01(0.89-1.14) | 0.900 |

Yellow and green colors indicate models with SVG azimuth tertiles (T1-T2-T3) and quartiles (Q1-Q2-Q3-Q4).

**Supplemental Table 7.**
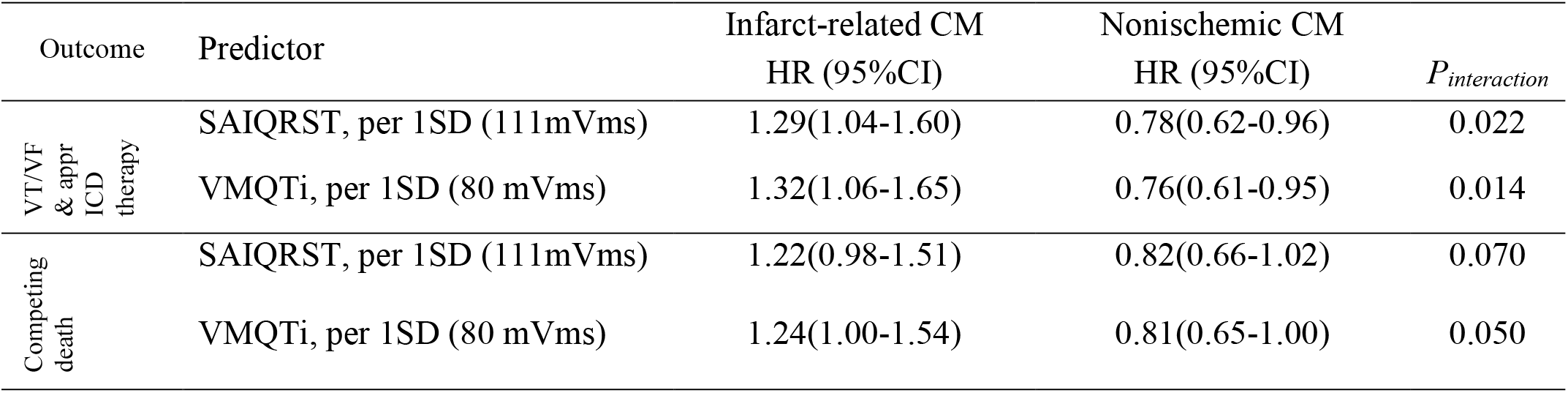
Adjusted (model 3) cause-specific hazard of two competing outcomes in ICM and NICM subgroups.

